# Funmap: integrating high-dimensional functional annotations to improve fine-mapping

**DOI:** 10.1101/2024.06.25.24309459

**Authors:** Yuekai Li, Jiashun Xiao, Jingsi Ming, Yicheng Zeng, Mingxuan Cai

## Abstract

Fine-mapping aims to prioritize causal variants underlying complex traits by accounting for the linkage disequilibrium of GWAS risk locus. The expanding resources of functional annotations serve as auxiliary evidence to improve the power of fine-mapping. However, existing fine-mapping methods tend to generate many false positive results when integrating a large number of annotations. In this study, we propose a unified method to integrate high-dimensional functional annotations with fine-mapping (Funmap). Funmap can effectively improve the power of fine-mapping by borrowing information from hundreds of functional annotations. Meanwhile, it relates the annotation to the causal probability with a random effects model that avoids the over-fitting issue, thereby producing a well-controlled false positive rate. Paired with a fast algorithm, Funmap enables scalable integration of a large number of annotations to facilitate prioritizing multiple causal SNPs. Our simulations demonstrate that Funmap is the only method that produces well-calibrated FDR under the setting of high-dimensional annotations while achieving better or comparable power gains as compared to existing methods. By integrating GWASs of 4 lipid traits with 187 functional annotations, Funmap consistently identified more variants that can be replicated in an independent cohort, achieving 15.5% − 26.2% improvement over the runner-up in terms of replication rate.

## Introduction

Genome-wide association studies (GWASs) have successfully identified hundreds of thousands of single nucleotide polymorphisms (SNPs) that are statistically associated with complex human traits [1]. However, because the true causal variants may be correlated with many non-causal variants located in the proximal region [2] due to linkage disequilibrium (LD), it remains very difficult to distinguish the causal SNPs from non-causal ones in GWAS discoveries. To address this issue, fine-mapping [3] is employed to prioritize the set of variants that are most likely to be biologically associated with a target trait while accounting for the local LD pattern within each genetic locus identified by GWAS. By offering a set of putative causal SNPs that are potentially responsible for panthology of complex human diseases, fine-mapping outputs can serve as valuable resources for in-depth exploration to interpret disease etiology.

A number of fine-mapping methods have been developed to prioritize causal variants based on GWAS data. Early methods for fine-mapping [4, 5] rely on an exhaustive search for combinations of causal SNPs, making them computationally inefficient in identifying multiple causal SNPs. Later, some methods employ approximate algorithms to reduce the computational cost of searching causal variants [6, 7, 8, 9]. More recently, SuSiE [10, 11] proposes an efficient fine-mapping framework by decomposing causal signals into a sum of single causal effects. Despite the great advances in fine-mapping, it remains a major challenge to reliably prioritize causal SNPs when they are in strong LD with non-causal ones.

Fortunately, functional annotations can serve as auxiliary information to inform the prioritization of causal SNPs. This is granted by the fact that biologically important SNPs are more enriched within functionally important annotations across the genome. For example, a fine-mapping analysis across 14 traits reported that putative causal SNPs were significantly enriched in functional regions, including nonsynonymous regions, conserved regions, and multiple cell-type-specific regulatory regions [12].

An expanding amount of annotation resources are becoming available to facilitate genetic studies. The Encyclopedia of DNA Elements (ENCODE) project [13] has generated a comprehensive mapping between functional elements and variants covering 80% of the genome, including open chromatin sites, histone mark enriched regions, and transcription factor binding regions, etc. The NIH Roadmap Epigenomics Mapping Consortium [14] has established high-quality, genome-wide maps of epigenomic regulatory elements across hundreds of human cell types and tissues, such as key histone modifications, chromatin accessibility, DNA methylation, and mRNA expression. Although the rich annotation resources hold promise to enhance fine-mapping, it requires handling high-dimensional annotation data, which hampers the effective integration of functional annotations. The difficulties are twofold. First, a large number of model parameters is required to characterize the relationship between annotations and the causal status of candidate SNPs. These parameters need to be estimated with GWAS data of only a few thousand correlated SNPs from a single genomic region, making the parameter estimation highly unreliable, thereby producing false positive results. Second, when the number of annotations grows large, it is computationally challenging to simultaneously search for multiple causal variants and estimate model parameters of high-dimensional annotations.

Much effort has been devoted to integrating functional annotations with GWAS data for fine-mapping. fastPAINTOR [15] can integrate multiple functional annotations with summary statistics by leveraging the approach of Markov chain Monte Carlo (MCMC). However, due to the lack of feature selection to filter out unrelated annotations, fastPAINTOR may incur a high false discovery rate (FDR) when dealing with high-dimensional functional annotations. To address this issue, PolyFun [12] proposes to first estimate prior weights based on functional annotations and GWAS summary statistics, and then perform other fine-mapping methods with these estimated prior weights. Because the two-step design of PolyFun does not maximize the joint-likelihood function, it usually has suboptimal statistical power. CARMA [16] attempts to improve existing methods by incorporating high-dimensional functional annotations via a penalized logistic regression, allowing for more reliable integration of summary data and high-dimensional functional annotations. However, CARMA’s algorithm alternates between a sampling procedure to explore the posterior distributions and fitting an elastic net to estimate annotation weights that involve a cross-validation step, making its time complexity relatively high as the number of candidate SNPs increases. Very recently, SparsePro [17] extends SuSiE by allowing the prior causal probabilities to be linked to binary functional annotations. Nevertheless, similar to PolyFun, the process of computing prior weights and fitting the SuSiE model in SparsePro is separated, and the functional annotations used in SparsePro are restricted to binary types.

In this paper, we propose a unified method to integrate high-dimensional <u>fun</u>ctional annotations with GWAS data to improve fine-<u>map</u>ping (Funmap). The success of Funmap relies on its three unique features. First, it effectively improves the power of fine-mapping by fully utilizing the information of high-dimensional functional annotations. Second, it relates the high-dimensional functional annotation to the prior causal probability with a random effects model that avoids the over-fitting issue, thereby producing a well-controlled FDR. Third, paired with a fast variational Bayes algorithm, Funmap enables scalable integration of a large number of annotations to facilitate prioritizing multiple causal SNPs. With comprehensive simulation studies, we show that Funmap is the only method that produces well-calibrated FDR under the setting of high-dimensional annotations while achieving better or comparable power gains as compared to existing fine-mapping methods. We applied Funmap to prioritize causal SNPs of 4 lipid traits by integrating their GWAS data with 187 functional annotations. Our results suggest that Funmap not only boosts the statistical power by fully leveraging the auxiliary evidence of function annotations but also substantially improves the reproducibility of putative causal SNPs, indicating its effectiveness in reducing false positives.

## Materials and methods

### The Funmap model

We begin the formulation of Funmap with the individual level GWAS data. Consider the GWAS dataset {**X, y**}, where **X** ∈ ℝ^*n*×*p*^ is the genotype matrix of *n* individuals and *p* SNPs of the target region, and **y** ∈ ℝ^*n*^ is the phenotype vector. Without loss of generality, we assume that each column of **X** and **y** has been standardized to have zero mean and unit variance. We also assume that the covariates, such as gender, age, and genotype principal components, have been properly adjusted following our previous works [18, 19]. We relate the phenotype **y** to genotypes **X** with the following linear model:

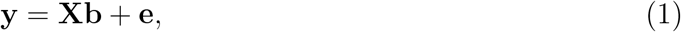

where **b** ∈ ℝ^*p*^ is the sparse vector of SNP effect sizes, the independent noise **e** ∼ 𝒩(**0**, *σ*^2^**I**_*n*_), and **I**_*n*_ is the *n* by *n* identity matrix. To identify the non-zero entries of **b**, we consider the following sum-of-single-effects [10] structure:

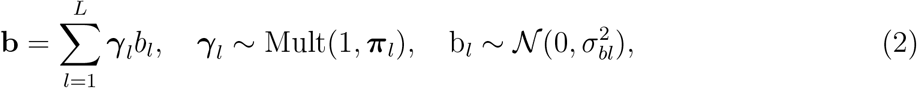

where ***γ***_*l*_ = [*γ*_*l*1_, …, *γ*_*lp*_]^*T*^ ∈ {0, 1}^*p*^ is a binary vector with *γ*_*lj*_ = 1 indicating the *l*-th causal signal is attributed to the *j*-th SNP, *b*_*l*_ is the effect size of of the *l*-th causal signal, and ***π***_*l*_ = [*π*_*l*1_, …, *π*_*lp*_]^*T*^ ∈ [0, 1]^*p*^ is the vector of prior causal probabilities with 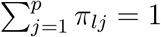.

Suppose that besides the GWAS data, we also collected the functional annotations of the target SNPs. Let **A** = [**A**_1_, …, **A**_*p*_]^*T*^ ∈ ℝ^*p*×*m*^ be the matrix collecting *m* annotations of the *p* SNPs. With the availability of rich functional annotations, *m* is becoming increasingly large (e.g., *m* ≥ 100). To effectively incorporate the functional annotation as auxiliary evidence for causal SNP prioritization, we link the prior probability ***π***_*l*_ to SNP annotations with the following softmax model with random effects:

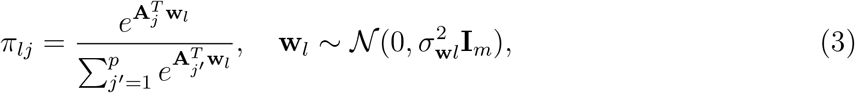

where **w**_*l*_ ∈ ℝ^*m*^ represents the random effects vector of annotations on the causal probability of the *l*-th single-effect component. Unlike previous methods [17, 15] that assume a fixed annotation effect shared across single-effect components, we propose a component-specific random effects assumption on the annotation weights **w**_*l*_. This assumption has two salient properties. First, it allows the annotation weights to vary across causal signals, which better characterizes the real genetic architecture. Second, built upon a random effects model, when dealing with high-dimensional functional annotation **A**, Funmap can adaptively estimate the annotation weights **w**_*l*_ from the data while shrinking the those of redundant annotations to avoid over-fitting, thereby effectively reduce false positive results.

Under Equations (1-3), we denote the model parameters as 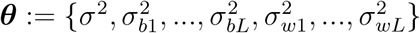 and the collections of random variables 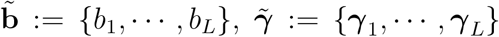, and 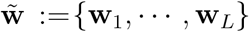. The logarithm of the marginal likelihood is given as 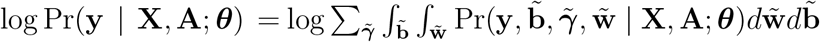 By maximizing the log-likelihood, we aim to obtain parameter estimates 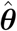 and prioritize the causal SNPs by using the posterior probability:

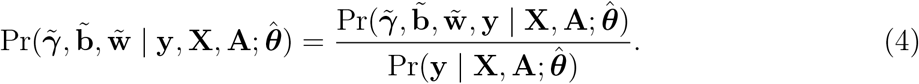

### Algorithm and parameter estimation

The major chellenge to evaluate Equation (4) is the intractable marginal likelihood on the denominator. First, it is difficult to exactly evaluate the integration of 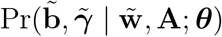 due to the softmax function. Second, the sum-of-single-effects assumption makes it intractable to integrate over 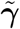. To address the above issues, we develop a variational expectation-maximization (EM) algorithm that can efficiently estimate model parameters and approximate the posterior distribution.

We first deal with the intractability resulting from the softmax function. Here, we consider the following bound based on double majorization [20]:

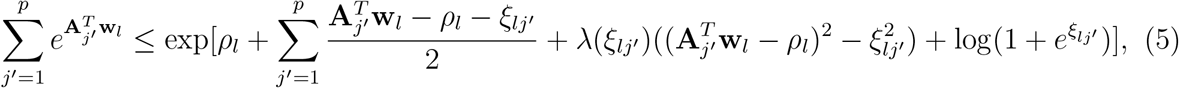

where 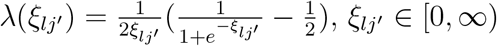, and *ρ* ∈ ℝ. Clearly, the right-hand side of this inequality is in the exponentiated quadratic form. Therefore, applying this inequality to the denominator of the softmax function (3) leads to a tractable bound 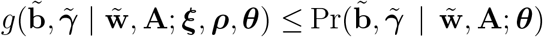, where ***ξ*** = [*ξ*]_*lj*_ ∈ [0, ∞)^*L*×*p*^ and ***ρ*** = [*ρ*_1_, …, *ρ*_*L*_]^*T*^ ∈ ℝ^*L*^ are variational parameters that can be estimated from the data (see details in Supplementary Note). Let **Θ** = {***θ, ξ, ρ***}. We can obtain a lower bound of complete-data likelihood as:

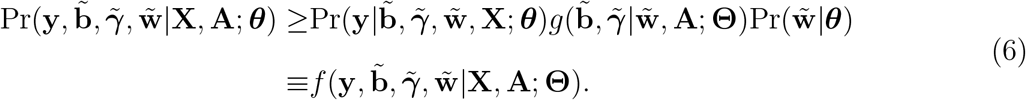

Based on the above bound, we further derive a lower bound of the logarithm of marginal likelihood

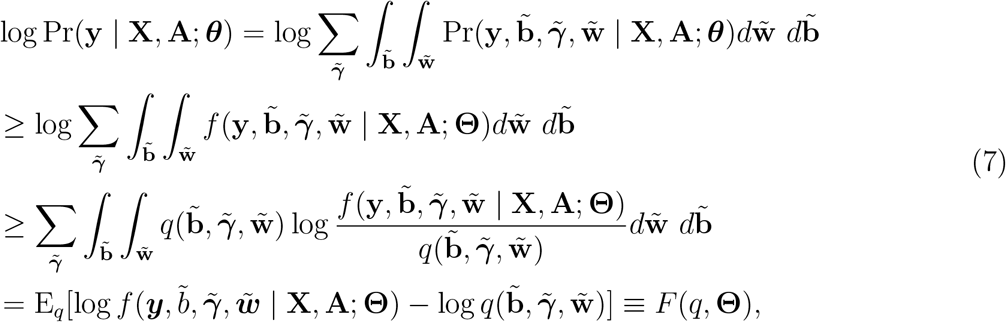

where the first i nequality f ollows t he d ouble m ajorization b ound, t he s econd i nequality is granted by Jensen’s inequality, and 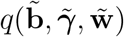 is an approximation of the exact posterior (4). To analytically evaluate the lower bound, we introduce the following factorizable mean-field formulation assumption:

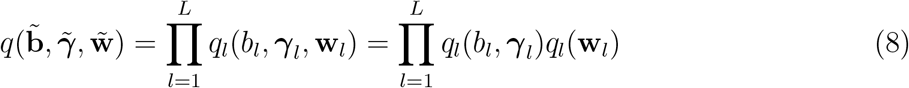

where *q*_*l*_(*b*_*l*_, ***γ***_*l*_) = *q*(*b*_*l*_ | ***γ***_*l*_)*q*(***γ***_*l*_). The above approximation can be obtained in closed form, which allows the lower bound to be analytically evaluated. Besides, by taking advantage of the sum-of-single-effects decomposition, this mean-field assumption inherits the property of SuSiE that only requires the *L* causal signals to be independent, relaxing the assumptions in traditional variational approximations [21, 22]. We develop a variational EM algorithm that can efficiently estimate ***θ*** and evaluate 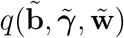 by iteratively maximizing the lower bound (7) under assumption (8).

To improve the convergence of the variational EM algorithm, we design a 3-stage model fitting process with warm-starts. This is built upon the knowledge that Funmap covers the SuSiE model as a special case when all the annotation effects 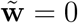. In the first stage, we fit the SuSiE model using our Funmap implementations without incorporating annotations, obtaining an initial estimate of parameters 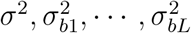 and the posterior distribution 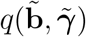. Then, we use these estimates to initialize the second stage where the variational EM algorithm is executed to evaluate the posterior distributions of 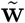. In this stage, we fix 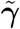 at its posterior mean obtained in the first stage to produce rough estimates of annotation-related parameters 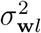 and 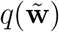. Finally, we use the estimated parameters from the second stage as the initial values to run the variational EM algorithm with the full Funmap model.

The Funmap fitting procedure possesses several appealing computational and practical convenience. Computationally, since the three fitting stages are constructed under a unified variational framework, the lower bound is guaranteed to increase at each stage, enabling a more stable convergence of the Funmap algorithm [23]. Practically, it avoids the need to specify the number of causal variants. Specifically, when the number of causal SNPs is unknown, we can simply set *L* to a reasonably large number. The excessive components of Funmap will be assigned small posterior probabilities. This property can be attributed to two facts. Firstly, at the first stage, excessive components will be broadly assigned to all SNPs across the locus due to the high uncertainty in allocating their causal effects [10, 24]. Secondly and importantly, the warm-start procedure can prevent the excessive components from being enlarged by over-fitting the high-dimensional annotations. We provide the details of our algorithm in the Supplementary note.

### Funmap with GWAS summary data

While the variational EM algorithm proposed above is for individual-level genotype **X** and phenotype **y**, it can be easily extended to using only GWAS summary data as input. Let us consider the *z*-scores obtained from marginal regressions:

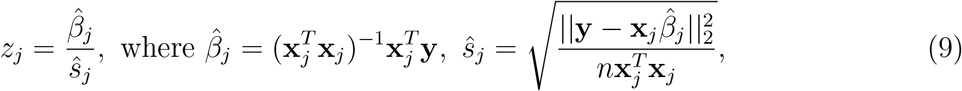

and **x**_*j*_ ∈ ℝ^*p*^ is the *j*-th column of **X**. We first note that the likelihood and its lower bound (7) depend on the GWAS data {**X, y**} only through the sufficient statistics **X**^*T*^ **X, X**^*T*^ **y** and **y**^*T*^ **y**. Since both **X** and **y** have been standardized, we can replace the sufficient statistics with *z*-scores and LD matrix **R** [10] with the following relationships:

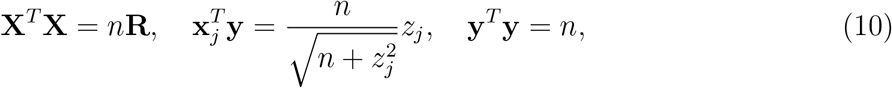

where **R** can be computed with genotypes from a subset of GWAS samples or from a reference panel of similar ancestry background. In our software implementation, all the operations are based on sufficient statistics, allowing us to accommodate both individual-level and summary-level GWAS data.

### Identification of causal SNPs

After the convergence of variational EM algorithm, we can obtain the approximate component-specific posterior probabilities *q*(***γ***_*l*_), where *q*(***γ***_*lj*_ = 1) represents the probability that *j*-th SNP is responsible for the *l*-th causal signal. With this information, we can compute the posterior inclusion probability of the *j*-th SNP as 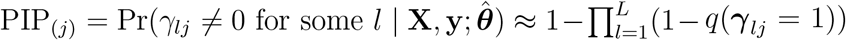. Then, the local FDR for each SNP can be calculated as fdr_(*j*)_ = 1 − PIP_(*j*)_, *j* = 1, *…*, *p*. To control the global FDR [25], we sort the local FDRs in ascending order. We denote the *i*-th sorted local FDR as fdr_(*i*)_ and compute the global FDR as 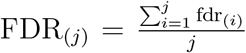.. If FDR_(*j*)_ is lower than a given FDR threshold *η*, we consider the SNP as a putative causal SNP.

## Results

### Simulation

We conducted comprehensive simulations to evaluate the performance of Funmap using real genotypes obtained from the UK Biobank (UKBB) project [26]. First, we obtained genotypes of UKBB White British individuals with sample sizes *n* = 20, 000 and 50, 000. To mimic the realistic genetic architectures of disease-related loci, we considered 10 risk regions identified in GWAS as associated with breast cancer [27], where each region comprises approximately *p* = 700 ∼ 4, 200 variants (Supplementary Table 1). The functional annotations *A*_*jk*_ of these SNPs were sampled from a standard normal distribution for *j* = 1…, *p* and *k* = 1, …, *m*. We varied the number of annotations *m* among {20, 50, 100} to cover the cases from low to high dimensional settings. Then, we constructed the effects of functional annotations **w** ∈ ℝ^*m*^ with *w*_*k*_ ∼ 𝒩(0, 0.01) for *k* = 1, …, *m*, where each annotation only has a small impact on the causal probability. Next, we simulated the causal probabilities with the softmax function 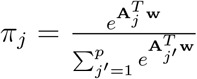. We selected *L*_0_ SNPs as the true causal SNP which has the highest values of *π*_*j*_ and pairwise correlation less than 0.1 following previous studies [17, 16]. We considered *L*_0_ = 2 and *L*_0_ = 3 to focus on scenarios where a locus harbours multiple causal variants. Given the causal status, we followed previous studies [10] to sample the effect sizes of causal SNPs 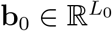 from 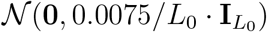, where 0.0075 is the heritability contributed by the target region. With the standardized genotype matrix of causal SNPs 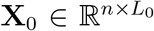, we generated the phenotype values with **y** = **X**_0_**b**_0_ + **e**, where 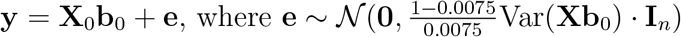. Finally, we performed univariate linear regressions on each SNP to obtain their *z*-scores. For each setting, we repeated the experiment for 50 times across the 10 regions, yielding 500 replicates. All the setting parameters and results are in Supplementary Figures 1–30.

In our simulation analysis, we benchmarked the performance of Funmap by comparing it with 5 representative fine-mapping methods. For methods that do not use functional information, we considered SuSiE [10], CARMA [16], and PAINTOR [15]. For methods that use functional information, we considered the extensions of CARMA and PAINTOR for integrating functional annotations, denoted as ‘CARMA+anno’ [16] and ‘PAINTOR+anno’[15], respectively. The number of components *L* for SuSiE and Funmap was set to 10, with no more than 100 iterations per stage. The expected number of causal SNPs (*η*) for CARMA and CARMA+anno was also set to 10, and the maximum number of iterations was set to 10 for both outer and inner loops. For PAINTOR and PAINTOR+anno, we used the fastPAINTOR implemented in the PAINTOR software v3.0 by adding the ‘-mcmc’ flag.

We first assessed the FDR calibration of the compared methods. Specifically, we calculated the global FDR defined and identified a set of putative causal SNPs with an expected FDR threshold. Given the putative causal SNPs, the empirical FDR can be computed as 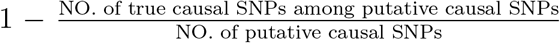. The eFDR of a well-calibrated fine-mapping method should be close to the expected FDR level. Figure 1a and 1b show the eFDR against expected FDR when *n* = 50, 000 and *L*_0_ = 2. Clearly, all methods without annotation had well-controlled FDR across different thresholds. Funmap consistently achieved satisfactory performance across different number of annotations, suggesting its ability to handle high-dimensional annotations through the random effects model. By contrast, due to the fixed effect assumption on annotation weights, PAINTOR+anno exhibited severely inflated FDR, which was exacerbated as the number of annotations increased. Although CARMA+anno could also regularize the annotation weights by fitting an elastic net, its FDR was still inflated when there was a large number of annotations (*m* ≥ 50). This may be attributed to the lack of warm-start procedure in CARMA’s algorithm, which leads to local optimal solutions. To illustrate the differences between the performance of compared methods more intuitively, we visualized the results of one dataset from our simulation experiments with Manhattan plots (Figure 1c). We can see that in this example, the two causal SNPs are much more significant than those of the surrounding SNPs due to the weak LD. Given this evidence, it is relatively easy to prioritize the causal SNPs in this dataset. Indeed, the causal SNPs could be reliably identified without the functional information. However, after incorporating functional annotations, CARMA+anno and PAINTOR+anno produced a number of false positive results. PINTOR+anno created 4 false positive signals with PIP> 0.9. CARMA+anno also incorrectly assigns a high PIP (> 0.8) to three non-causal SNPs. By contrast, Funmap effectively avoided false positive findings and yielded results similar to SuSiE. This observation implies the effectiveness of using Funmap’s random effects model to adaptively incorporate functional information in high-dimensional settings.

**Figure 1:**
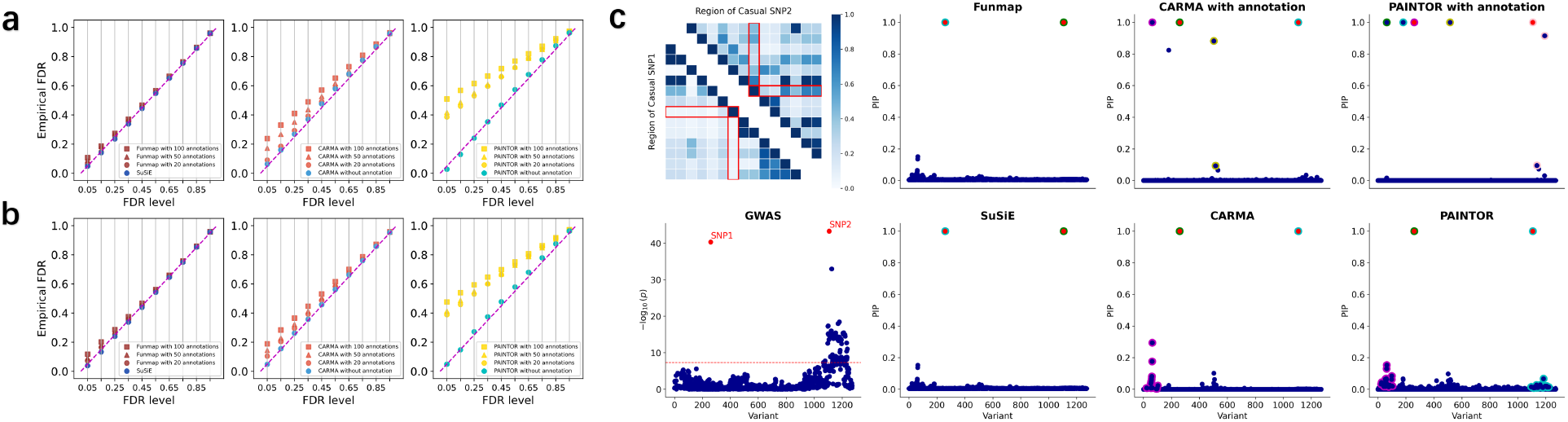
Comparison of FDR control in simulation studies. **a**-**b**. Calibration of FDR with *n* = 50, 000, *m* = 100, while the number of causal SNPs is set to *L*_0_ = 2 (**a**) and *L*_0_ = 3 (**b**). Results are summarized from 500 replications across 10 regions. **c**. An illustrative example generated by simulation. The first column shows the absolute correlation among the two candidate causal SNPs and their neighboring SNPs and the Manhattan plot. The second to fourth columns show the PIP obtained by with compared methods. Red dots represent causal SNPs. Circles in the same color represent SNPs included in the level-95% credible sets of a causal signal.

Next, we considered a set of common PIP thresholds {0.90, 0.95, 0.99} to evaluate the statistical power of compared methods. As shown in Figure 2a and Figure 2b, by incorporating functional annotation information, the statistical power of Funmap, CARMA+anno, and PAINTOR+anno outperformed their counterparts without annotations. The statistical power of Funmap was higher than CARMA+anno when stringent PIP thresholds (PIP>0.99 and PIP>0.95) were applied and comparable to CARMA+anno under a less stringent PIP threshold (PIP4>0.9). Although PAINTOR+anno produced the highest power in most settings, it is worth noting that its FDR can be strongly inflated when then number of annotations is large, making it difficult to replicate and interpret the discoveries. A concrete example with *m* = 100 and *L*_0_ = 2 is given in Figure 2c, where the two causal SNPs are in strong LD with neighboring SNPs. SuSiE and CARMA produced a PIP of around 0.8 for the causal SNPs, while PAINTOR’s PIP failed to exceed 0.4. Despite the strong LD, Funmap successfully elevated the PIP of causal SNPs to 1.0 by utilizing the functional information. In contrast, CARMA+anno remained largely unchanged and PAINTOR+anno incorrectly assigned high PIP to a non-causal SNP, yielding a false positive result. This example indicates that even in the presence of strong LD, traditional methods can have limited power, whereas Funmap can still effectively improve statistical power by leveraging the functional information while controlling FDR. To gain further insight into the difference in fine-mapping performance, we contrasted the PIP obtained by Funmap against those obtained by other methods (Figure 3a and b). It is clear that the PIP of causal variants produced by Funmap was substantially larger than those produced by SuSiE and CARMA+anno, indicating Funmap’s better ability to pinpoint the causal SNPs. As a comparison, PAINTOR+anno identified more causal SNPs with a cost of producing many false positives. This is in line with our observation of inflated FDR yielded by PAINTOR+anno. Besides the PIP for SNP-level inference, Funmap also provides a set of candidate SNPs for each causal signal, denoted as ‘credible set’. A level-*δ* credible set includes a subset of SNPs that are responsible for a single-effect component with a joint probability of *δ*. Given a coverage level *δ*, a smaller credible set indicates a lower uncertainty to assign the causal signal and hence higher resolution of fine-mapping. We summarized the size of level-95% credible sets in Figure 3d. As expected, Funmap yielded the smallest credible sets, suggesting its higher resolution.

**Figure 2:**
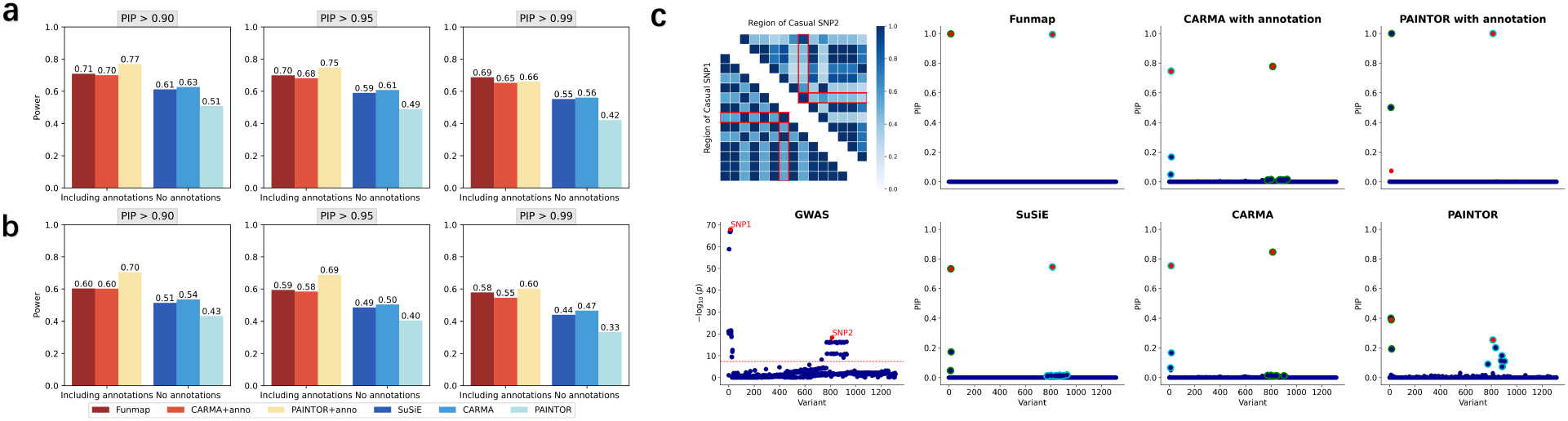
Comparison of statistical power in simulation studies. **a**-**b**. Statistical power of compared methods with *n* = 50, 000, *m* = 100 while the number of causal SNPs is set to *L*_0_ = 2 (**a**) and *L*_0_ = 3 (**b**). **c**. An illustrative example generated by simulation.

**Figure 3:**
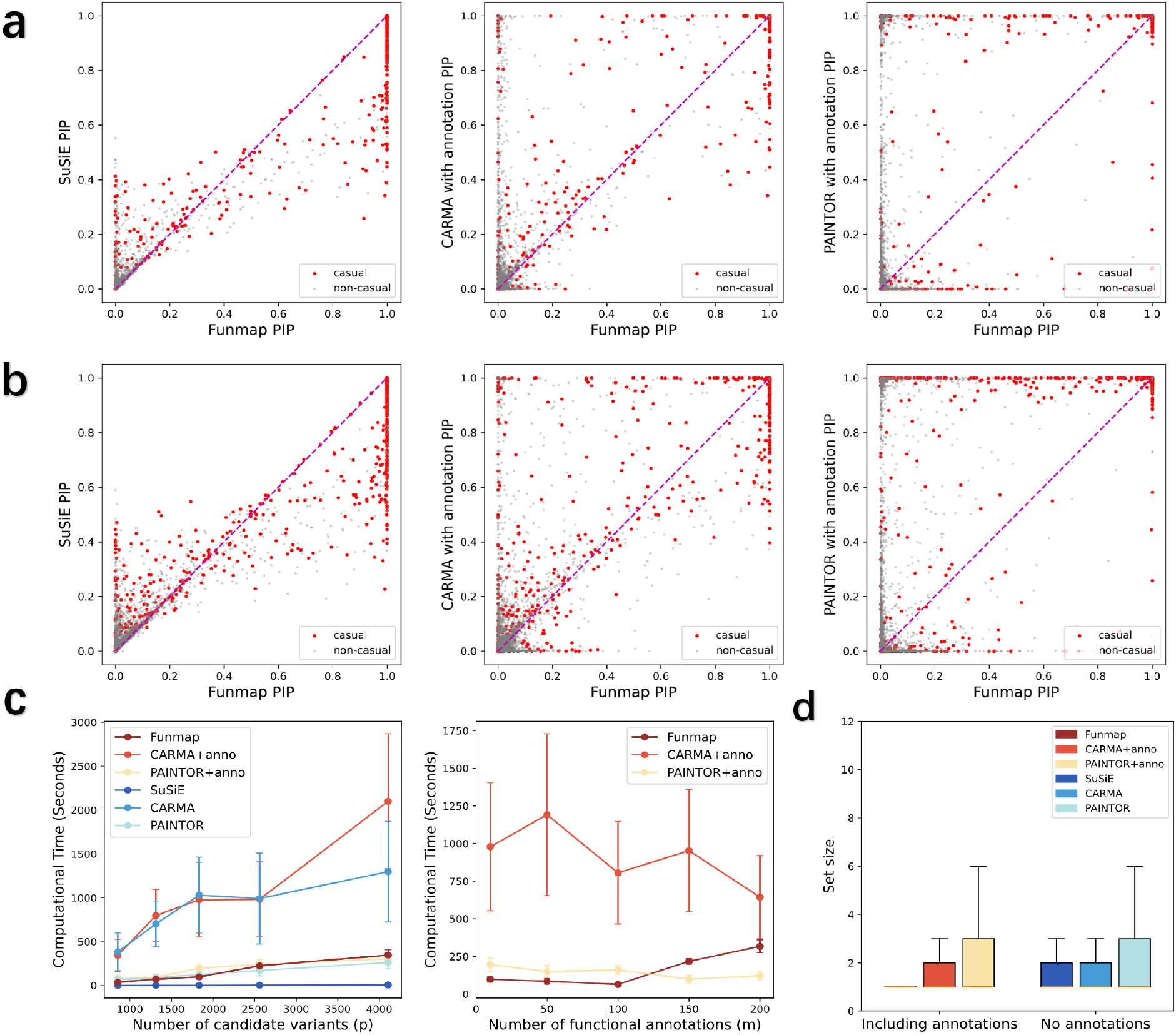
Comparison of PIP and CPU timings. **a**-**b**. Comparison of PIP between Funmap and SuSiE (left panel), CARMA+anno (middle panel), and PAINTOR+anno (right panel) with *n* = 50, 000, *m* = 100, while *L*_0_ is varied at 2 (**a**) and 3 (**b**). **c**. CPU timings are shown for increasing *p* with *m* = 100 (left panel) and increasing *m* with *p* = 1, 833. **d**. Boxplot displays the size of the 95% credible sets from the simulation results with *n* = 50, 000, *m* = 100, *L*_0_ ∈ {2, 3}.

To evaluate the computational efficiency of our model, we benchmarked the CPU time of compared methods by varying the number of variables and the number of functional annotations. We first focused on loci with number of SNPs *p* ∈ {854, 1313, 1833, 2561, 4107}, while fixing the number of functional annotations at *m* = 100. Figure 3b shows the CPU time of all compared methods. As we can observe, Funmap achieved a great computational efficiency among methods capable of integrating functional annotation, with a CPU time substantially faster than CARMA+anno. While SuSiE was the fastest method, it could not integrate high-dimensional annotations to improve fine-mapping. Figure 3c compares the running times under different numbers of functional annotations *m* ∈ {10, 50, 100, 150, 200} with the number of SNPs fixed at *p* = 1, 833. Overall, CARMA+anno had the largest computational overhead, with an average CPU time ranging between 10 and 16 minutes. Funmap achieved the least computing time when *m* ≤ 100 and only had a marginal increase when *m* exceeds 150. Although PAINTOR+anno had CPU times comparable Funmap, it is not suitable to integrate high-dimensional annotations due to inflated FDR.

To assess the robustness of Funmap when our assumption of annotation effects is violated, we conducted a series of additional simulations that consider the sparse annotation weights. Specifically, we generated the causal probabilities by setting half of the annotation weights to zero. Other simulation settings remained unchanged. As shown in Supplementary Figures 31-48, Funmap performed reasonably well in FDR control while improving statistical power when the distribution of annotation weights was misspecified.

### Real data analysis

We applied Funmap to identify causal SNPs for four lipid-related traits including high-density lipoprotein (HDL), low-density lipoprotein (LDL), triglycerides (TG), and total cholesterol (TC). The GWAS summary data were collected from 315,133 UKBB individuals of European ancestry. We obtained 187 functional annotations from the Baseline-LF v2.2.UKB annotations [28, 12], with details given in the Supplementary note. To construct candidate genomic regions for fine-mapping, we first identified genome-wide significant associations (p-values *<* 5 × 10−8) for each trait. Then, we extracted a 1-Mbp window centering at each genome-wide significant SNP. In total, we obtained 864 genomic regions (190-347 per trait) with 434-8646 SNPs per region across the four traits. To avoid spurious results as reported by previous studies [12], we excluded the region of major histocompatibility complex (25.5 Mbp–33.5 Mbp in chromosome 6) and two regions with long-range LD (8 Mbp–12 Mbp in chromosome 8 and 46 Mbp–57 Mbp in chromosome 11). During the fine-mapping process, we considered one genomic region at a time, using the GWAS marginal *z*-scores, in-sample LD correlation matrices, and the functional annotations of local SNPs as inputs for all methods. We recorded the PIPs and credible sets for each genomic region. We used the same parameter settings as those used in simulation studies for Funmap, SuSiE, and PAINTOR. For CARMA, we set the maximum number of iterations to 4 for both inner and outer loops.

We summarized the putative causal SNPs in Supplementary Table 2. By integrating functional annotations, Funmap successfully identified more SNPs than SuSiE, PAINTOR, and CARMA across different PIP thresholds. Although PAINTOR+anno and CARMA+anno reported more causal SNPs than Funmap, our simulation analyses suggest that their discoveries may be unreliable due to the large number of annotations. To assess the credibility of our fine-mapping results, we extracted the set of SNPs that were reported as causal by each functionally informed method but not by SuSiE with a PIP threshold of 0.95. Then, we evaluated the replication rates of these newly identified SNPs using independent multi-ancestry GWASs from the Global Lipids Genetics Consortium (GLGC) [29], which comprised up to 900K samples across the four traits. The credible sets in this replication cohort were computed by SuSiE using meta-analyzed multi-ancestry data, yielding a highly reliable set of putative causal SNPs. Therefore, we can use the GLGC data as a high-quality resource to validate the fine-mapping results in our UKBB analysis. We evaluated the replication rates with two quantities: the proportion of new discoveries that were genome-wide significant (*p*-value*<* 5 × 10^−8^) and the proportion of new discoveries included in the credible sets in GLGC cohort. As summarized in Figure 4, Funmap consistently yielded the highest replication rates across the four traits, with > 50% new discoveries successfully replicated in HDL, LDL, and TC in terms of genome-wide significance. Among the three methods that integrate functional annotations, PAINTOR+anno detected 1,110 new associations with HDL that were not reported by SuSiE. However, only 17.8% (198*/*1110) were genome-wide significant, and 3.5% (39*/*1110) were included in the credible sets of GLGC cohort. Although CARMA+anno had better replication performance than PAINTOR+anno, only 31.5% (198*/*629) new discoveries were genome-wide significant in GLGC, meaning that more than half of its new discoveries could not be replicated. The low replication rates suggest that many new discoveries could be false positives due to the integration of high-dimensional annotations. By introducing the random effects model, Funmap substantially improved the replication rates. For example, in terms of genome-wide significance, 53.3%(202*/*379) new SNPs reported by Funmap were successfully replicated. Besides, 16.7% (34*/*203) of Funmap’s new discoveries were included in the GLGC credible sets, which was 2.5 times higher than CARMA+anno and 4.8 times higher than PAINTOR+anno. It is worth noting that the number of Funmap’s new discoveries replicated by GLGC credible sets was consistently higher, implying a greater statistical power. Meanwhile, the number of non-replicated SNPs was substantially reduced, indicating that many false positives were excluded. This pattern was consistent across the four traits. Therefore, the higher replication rates of Funmap can be attributed to both improved power and reduced false positives by properly integrating the high-dimensional annotations.

**Figure 4:**
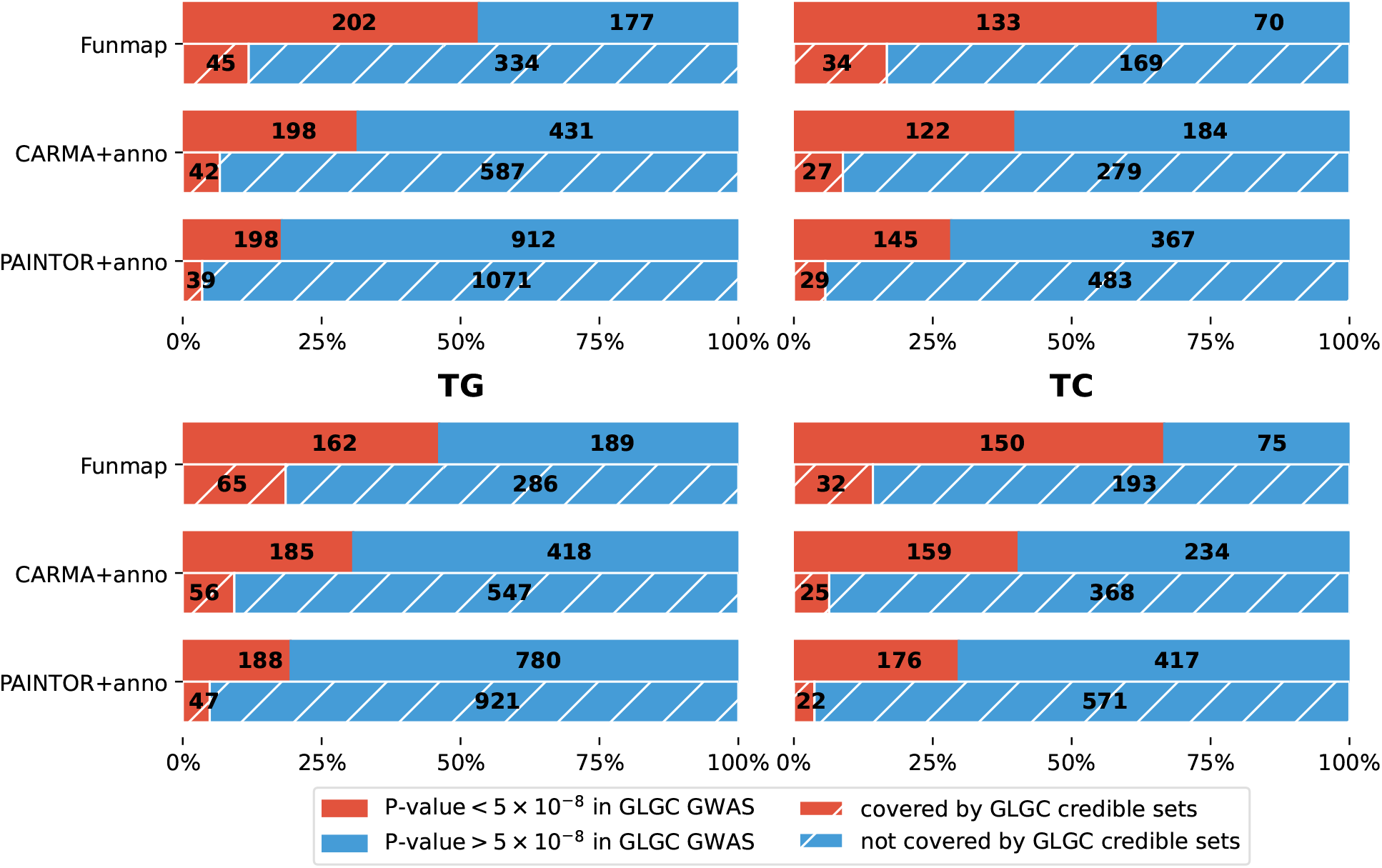
Replication analysis of Funmap, CARMA+anno, and PAINTOR+anno. Bar charts on the top shows the fraction and number of newly identified SNPs with *p*-value*<* 5 × 10^−8^ in the replication cohorts of GLGC GWAS. Bar charts on the bottom shows the fraction and number of newly identified SNPs that are included in the 95%-level credible sets generated from GLGC GWAS with SuSiE.

Funmap not only improved the replication rates but also enhanced the fine-mapping resolution in the lipid traits analysis. As observed in Figure 5a, Funmap created the smallest credible sets among all compared methods, with a median size of one across the four traits. This observation was consistent with our simulation studies. As an example, we focus on the locus 6-7Mb in Chromosome 8, which harbours the SNP rs2928176 fine-mapped for TC only by Funmap (PIP=1.0). This SNP locates on the 2Kb upstream of *AGPAT5*, encoding an integral membrane protein of the 1-acylglycerol-3-phosphate O-acyltransferase family [30]. This enzyme is responsible for converting lysophosphatidic acid to phosphatidic acid, serving as a critical step of de novo phospholipid biosynthesis. The SNP correlation heatmap (top left panel in Figure 5b) in this region shows that multiple SNPs around rs2928176 are in strong LD, making them highly significant with very similar *p*-values (bottom left panel in Figure 5b). Therefore, traditional fine-mapping methods lacking the ability to integrate functional information could not reliably prioritize the causal SNP in this region. Indeed, SuSiE produced a very large 95% credible set comprising 41 SNPs with the largest PIP only attained 0.217. CARMA and PAINTOR generated credible sets of similar sizes. By incorporating the functional information, Funmap uniquely identified rs2928617 as the causal SNP with *PIP* = 1, producing a high-resolution credible set that only includes rs2928617. By contrast, CARMA+anno yielded three credible sets with sizes of one and two, which was suspicious as there is no strong evidence of three independent causal signals in this region. While PAINTOR+anno elevated the highest PIP to 0.8 and slightly reduced the size of the credible set, it failed to include rs2928617. This example consolidates our conclusion of Funmap’s ability to improve power and resolution by incorporating high-dimensional functional annotations.

**Figure 5:**
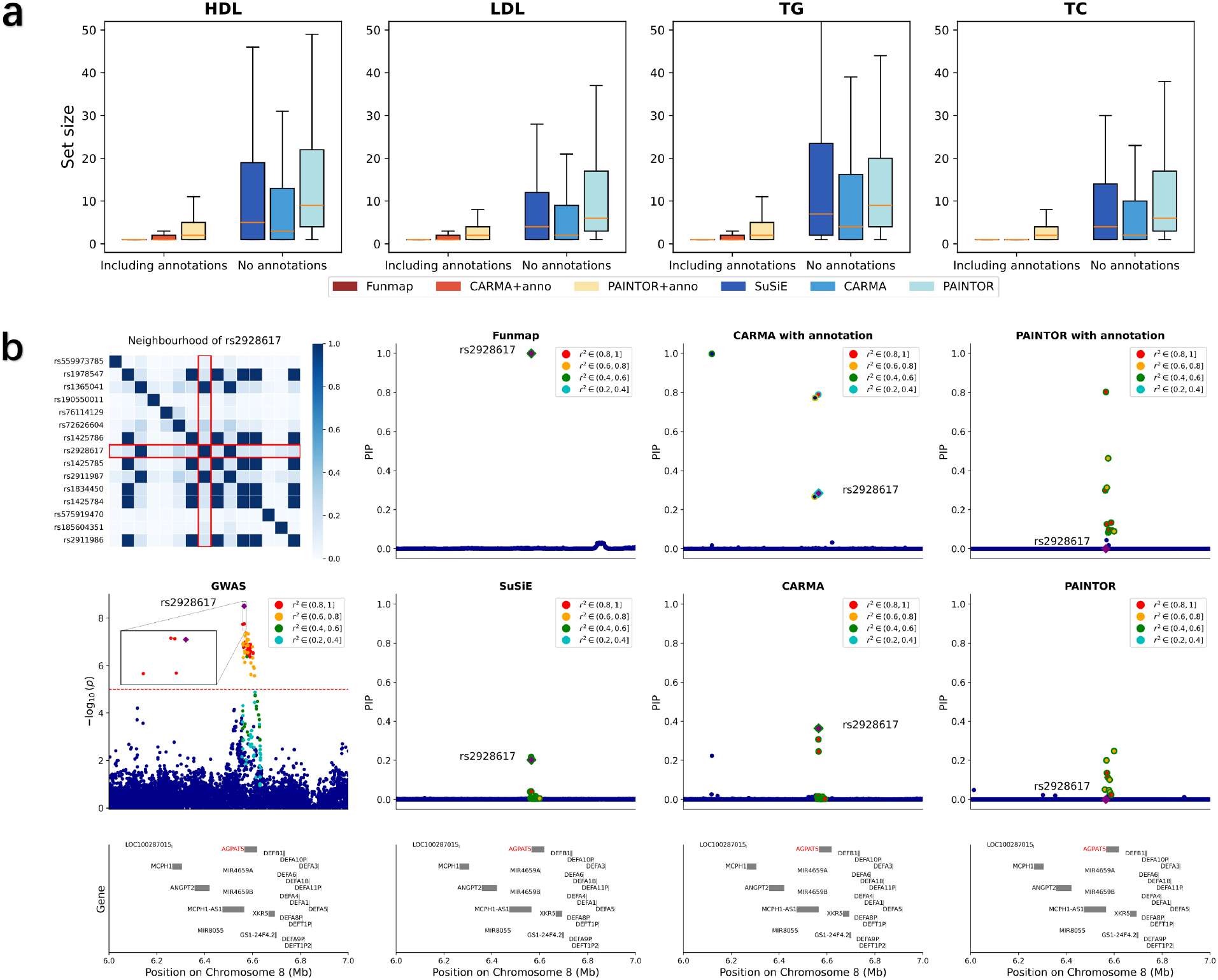
Comparison of credible set size and fine-mapping results from a region of TC GWAS. **a**. Box plots of credible set size across 4 lipid traits. **b**. Fine-mapping results of TC from locus 6 Mb–7 Mb in chromosome 8. The first column shows the heatmap of absolute correlation between rs2928617 and its neighboring SNPs and the Manhattan plot. The red dashed line represents 5 × 10^−8^. The second to fourth column show the PIP obtained by with compared methods. The purple square represents SNP rs2928617 and the color of the points represents the correlation between neighboring SNPs and rs2928617. Circles in the same color represent SNPs in the level-95% credible sets of a causal signal.

## Discussion and conclusion

In the paper, we introduced a novel and computationally efficient fine-mapping method, Funmap, to adaptively integrate functional information from a vast amount of SNP annotations. It not only boosts the statistical power of fine-mapping, but also produces well-controlled FDR when considering a large number of functional annotations. Funmap’s efficient algorithm allows it to simultaneously identify multiple causal signals while handling hundreds of annotations. Through comprehensive simulations, we showed that Funmap achieved greater statistical power and higher resolution while producing better-controlled FDR. We applied Funmap to identify causal SNPs for 4 lipid traits by integrating 187 functional annotations, yielding substantial power gains. Importantly, we showed that Funmap effectively avoided spurious results, identifying causal SNPs that could be better reproduced.

Although using GWAS summary statistics as input in fine-mapping offers practical convenience, it usually requires a population-match LD reference. Discrepancies such as inconsistent allele frequencies and ‘allele flip’ between summary data and LD reference can lead to biased PIP values [16, 11]. To avoid spurious discoveries, we have used in-sample LD for the fine-mapping analysis in this study. This strategy can minimize the chance of mis-matching. When the in-sample LD is not available, we recommend applying diagnostic methods, such as SLALOM [31], DENTIST [32], or the discrepancy detection module in CARMA [16], as a quality control step to exclude susceptible SNPs from analysis.

Our Funmap framework can be extended along several directions. First, the distinct recombination rates lead to very different LD patterns among global populations. Therefore, extending Funmap to leverage genetic diversity with cross-population GWASs may further improve fine-mapping power and resolution [24, 33, 34, 35]. Second, because the biological pathways underlying causal SNPs remain largely unknown, it is of particular interest to investigate the relevant annotations at the locus level. Funmap’s random effects model can be extended by introducing a sparse structure to map relevant annotations to each causal signal, yielding in-depth biological interpretation of causal SNPs [23]. Third, while our main analysis focuses on functional features related to genic region, allele frequency, and LD, publicly available resources at single-cell resolution are rapidly growing, serving as new data resources to annotate risk SNPs. Integration of fine-mapping with single-cell genomics, epigenomics, and proteomics data will be a promising direction to reveal the cellular contexts of causal signals [36].

## Supporting information

FunMap Supplementary

## Data Availability

All data produced are available online at:
https://nealelab.github.io/UKBB ldsc/index.html
https://alkesgroup.broadinstitute.org/UKBB LD.ThescATAC-seqdata
https://alkesgroup.broadinstitute.org/LDSCORE/baselineLD v2.1 annots
https://csg.sph.umich.edu/willer/public/glgclipids2021

https://github.com/LeeHITsz/Funmap

## References

[1] Emil Uffelmann, Qin Qin Huang, Nchangwi Syntia Munung, Jantina De Vries, Yukinori Okada, Alicia R Martin, Hilary C Martin, Tuuli Lappalainen, and Danielle Posthuma. Genome-wide association studies. Nature Reviews Methods Primers, 1(1):59, 2021.

[2] Montgomery Slatkin. Linkage disequilibrium—understanding the evolutionary past and mapping the medical future. Nature Reviews Genetics, 9(6):477–485, 2008.

[3] Daniel J Schaid, Wenan Chen, and Nicholas B Larson. From genome-wide associations to candidate causal variants by statistical fine-mapping. Nature Reviews Genetics, 19(8):491–504, 2018.

[4] Farhad Hormozdiari, Emrah Kostem, Eun Yong Kang, Bogdan Pasaniuc, and Eleazar Eskin. Identifying causal variants at loci with multiple signals of association. In Proceedings of the 5th ACM Conference on Bioinformatics, Computational Biology, and Health Informatics, pages 610–611, 2014.

[5] Gleb Kichaev, Wen-Yun Yang, Sara Lindstrom, Farhad Hormozdiari, Eleazar Eskin, Alkes L Price, Peter Kraft, and Bogdan Pasaniuc. Integrating functional data to prioritize causal variants in statistical fine-mapping studies. PLoS genetics, 10(10):e1004722, 2014.

[6] Wenan Chen, Beth R Larrabee, Inna G Ovsyannikova, Richard B Kennedy, Iana H Haralambieva, Gregory A Poland, and Daniel J Schaid. Fine mapping causal variants with an approximate bayesian method using marginal test statistics. Genetics, 200(3):719–736, 2015.

[7] Christian Benner, Chris CA Spencer, Aki S Havulinna, Veikko Salomaa, Samuli Ripatti, and Matti Pirinen. Finemap: efficient variable selection using summary data from genome-wide association studies. Bioinformatics, 32(10):1493–1501, 2016.

[8] Xiaoquan Wen, Yeji Lee, Francesca Luca, and Roger Pique-Regi. Efficient integrative multi-snp association analysis via deterministic approximation of posteriors. The American Journal of Human Genetics, 98(6):1114–1129, 2016.

[9] Yeji Lee, Francesca Luca, Roger Pique-Regi, and Xiaoquan Wen. Bayesian multi-snp genetic association analysis: control of fdr and use of summary statistics. BioRxiv, page 316471, 2018.

[10] Gao Wang, Abhishek Sarkar, Peter Carbonetto, and Matthew Stephens. A simple new approach to variable selection in regression, with application to genetic fine mapping. Journal of the Royal Statistical Society Series B: Statistical Methodology, 82(5):1273–1300, 2020.

[11] Yuxin Zou, Peter Carbonetto, Gao Wang, and Matthew Stephens. Fine-mapping from summary data with the “sum of single effects” model. PLoS Genetics, 18(7):e1010299, 2022.

[12] Omer Weissbrod, Farhad Hormozdiari, Christian Benner, Ran Cui, Jacob Ulirsch, Steven Gazal, Armin P Schoech, Bryce Van De Geijn, Yakir Reshef, Carla Márquez-Luna, et al. Functionally informed fine-mapping and polygenic localization of complex trait heritability. Nature genetics, 52(12):1355–1363, 2020.

[13] ENCODE Project Consortium et al. An integrated encyclopedia of dna elements in the human genome. Nature, 489(7414):57, 2012.

[14] Anshul Kundaje, Wouter Meuleman, Jason Ernst, Misha Bilenky, Angela Yen, Alireza Heravi-Moussavi, Pouya Kheradpour, Zhizhuo Zhang, Jianrong Wang, Michael J Ziller, et al. Integrative analysis of 111 reference human epigenomes. Nature, 518(7539):317–330, 2015.

[15] Gleb Kichaev, Megan Roytman, Ruth Johnson, Eleazar Eskin, Sara Lindstroem, Peter Kraft, and Bogdan Pasaniuc. Improved methods for multi-trait fine mapping of pleiotropic risk loci. Bioinformatics, 33(2):248–255, 2017.

[16] Zikun Yang, Chen Wang, Linxi Liu, Atlas Khan, Annie Lee, Badri Vardarajan, Richard Mayeux, Krzysztof Kiryluk, and Iuliana Ionita-Laza. Carma is a new bayesian model for fine-mapping in genome-wide association meta-analyses. Nature Genetics, 55(6):1057–1065, 2023.

[17] Wenmin Zhang, Hamed Najafabadi, and Yue Li. Sparsepro: An efficient fine-mapping method integrating summary statistics and functional annotations. PLoS genetics, 19(12):e1011104, 2023.

[18] Mingxuan Cai, Jiashun Xiao, Shunkang Zhang, Xiang Wan, Hongyu Zhao, Gang Chen, and Can Yang. A unified framework for cross-population trait prediction by leveraging the genetic correlation of polygenic traits. The American Journal of Human Genetics, 108(4):632–655, 2021.

[19] Jiashun Xiao, Mingxuan Cai, Xinyi Yu, Xianghong Hu, Gang Chen, Xiang Wan, and Can Yang. Leveraging the local genetic structure for trans-ancestry association mapping. The American Journal of Human Genetics, 109(7):1317–1337, 2022.

[20] Guillaume Bouchard. Efficient bounds for the softmax function and applications to approximate inference in hybrid models. In NIPS 2007 workshop for approximate Bayesian inference in continuous/hybrid systems, volume 6, 2007.

[21] Yongtao Guan and Matthew Stephens. Bayesian variable selection regression for genome-wide association studies and other large-scale problems. The Annals of Applied Statistics, 5(3):1780–1815, 2011.

[22] Peter Carbonetto and Matthew Stephens. Scalable variational inference for bayesian variable selection in regression, and its accuracy in genetic association studies. Bayesian analysis, 7(1):73–108, 2012.

[23] Jingsi Ming, Mingwei Dai, Mingxuan Cai, Xiang Wan, Jin Liu, and Can Yang. Lsmm: a statistical approach to integrating functional annotations with genome-wide association studies. Bioinformatics, 34(16):2788–2796, 2018.

[24] Mingxuan Cai, Zhiwei Wang, Jiashun Xiao, Xianghong Hu, Gang Chen, and Can Yang. Xmap: Cross-population fine-mapping by leveraging genetic diversity and accounting for confounding bias. Nature Communications, 14(1):6870, 2023.

[25] Bradley Efron. Size, power and false discovery rates. The Annals of Statistics, 35(4):1351 –1377, 2007.

[26] Clare Bycroft, Colin Freeman, Desislava Petkova, Gavin Band, Lloyd T Elliott, Kevin Sharp, Allan Motyer, Damjan Vukcevic, Olivier Delaneau, Jared O’Connell, et al. The uk biobank resource with deep phenotyping and genomic data. Nature, 562(7726):203–209, 2018.

[27] Laura Fachal, Hugues Aschard, Jonathan Beesley, Daniel R Barnes, Jamie Allen, Siddhartha Kar, Karen A Pooley, Joe Dennis, Kyriaki Michailidou, Constance Turman, et al. Fine-mapping of 150 breast cancer risk regions identifies 191 likely target genes. Nature genetics, 52(1):56–73, 2020.

[28] Alexander Gusev, S Hong Lee, Gosia Trynka, Hilary Finucane, Bjarni J Vilhjálmsson, Han Xu, Chongzhi Zang, Stephan Ripke, Brendan Bulik-Sullivan, Eli Stahl, et al. Partitioning heritability of regulatory and cell-type-specific variants across 11 common diseases. The American Journal of Human Genetics, 95(5):535–552, 2014.

[29] Sarah E Graham, Shoa L Clarke, Kuan-Han H Wu, Stavroula Kanoni, Greg JM Zajac, Shweta Ramdas, Ida Surakka, Ioanna Ntalla, Sailaja Vedantam, Thomas W Winkler, et al. The power of genetic diversity in genome-wide association studies of lipids. Nature, 600(7890):675–679, 2021.

[30] Anastasiya Strembitska, Gwenaël Labouèbe, Alexandre Picard, Xavier P Berney, David Tarussio, Maxime Jan, and Bernard Thorens. Lipid biosynthesis enzyme agpat5 in agrp-neurons is required for insulin-induced hypoglycemia sensing and glucagon secretion. Nature Communications, 13(1):5761, 2022.

[31] Masahiro Kanai, Roy Elzur, Wei Zhou, Kuan-Han H Wu, Humaira Rasheed, Kristin Tsuo, Jibril B Hirbo, Ying Wang, Arjun Bhattacharya, Huiling Zhao, et al. Meta-analysis fine-mapping is often miscalibrated at single-variant resolution. Cell Genomics, 2(12), 2022.

[32] Wenhan Chen, Yang Wu, Zhili Zheng, Ting Qi, Peter M Visscher, Zhihong Zhu, and Jian Yang. Improved analyses of gwas summary statistics by reducing data heterogeneity and errors. Nature Communications, 12(1):7117, 2021.

[33] Boran Gao and Xiang Zhou. Mesusie enables scalable and powerful multi-ancestry fine-mapping of causal variants in genome-wide association studies. Nature Genetics, pages 1–10, 2024.

[34] Kai Yuan, Ryan J Longchamps, Antonio F Pardiñas, Mingrui Yu, Tzu-Ting Chen, Shu-Chin Lin, Yu Chen, Max Lam, Ruize Liu, Yan Xia, et al. Fine-mapping across diverse ancestries drives the discovery of putative causal variants underlying human complex traits and diseases. medRxiv, 2023.

[35] Nathan LaPierre, Kodi Taraszka, Helen Huang, Rosemary He, Farhad Hormozdiari, and Eleazar Eskin. Identifying causal variants by fine mapping across multiple studies. PLoS genetics, 17(9):e1009733, 2021.

[36] Fulong Yu, Liam D Cato, Chen Weng, L Alexander Liggett, Soyoung Jeon, Keren Xu, Charleston WK Chiang, Joseph L Wiemels, Jonathan S Weissman, Adam J de Smith, et al. Variant to function mapping at single-cell resolution through network propagation. Nature Biotechnology, 40(11):1644–1653, 2022.

