## Supplementary material for "Funmap: integrating high-dimensional functional annotations to improve fine-mapping": FunMap Supplementary

### Supplementary note and figures of “Funmap: integrating high-dimensional functional annotations to improve fine-mapping”

June 24, 2024

**4 Supplementry table 29**

**5 Code and data availability 30**

### 1 Details of the method

#### 1.1 Model setup

Consider the GWAS dataset $\{\mathbf{X}, \mathbf{y}\}$, where $\mathbf{X} \in \mathbb{R}^{n \times p}$ is the genotype matrix of $n$ individuals and $p$ SNPs of the target region, and $\mathbf{y} \in \mathbb{R}^n$ is the phenotype vector. Without loss of generality, we assume that each column of $\mathbf{X}$ and $\mathbf{y}$ has been standardized to have zero mean and unit variance. We also assume that the covariates, such as gender, age, and genotype principal components, have been properly adjusted. We relate the phenotype $\mathbf{y}$ to genotypes $\mathbf{X}$ with the following linear model:

$$\mathbf{y} = \mathbf{X}\mathbf{b} + \mathbf{e}, \tag{1}$$

where $\mathbf{b} \in \mathbb{R}^p$ is the sparse vector of SNP effect sizes, the independent noise $\mathbf{e} \sim \mathcal{N}(\mathbf{0}, \sigma^2 \mathbf{I}_n)$, and $\mathbf{I}_n$ is the $n$ by $n$ identity matrix. To identify the non-zero entries of $\mathbf{b}$, we consider the following sum-of-single-effects [1] structure:

$$\mathbf{b} = \sum_{l=1}^{L} \boldsymbol{\gamma}_l b_l, \quad \boldsymbol{\gamma}_l \sim \text{Mult}(1, \boldsymbol{\pi}_l), \quad \text{b}_l \sim \mathcal{N}(0, \sigma_{bl}^2), \tag{2}$$

where $\boldsymbol{\gamma}_l = [\gamma_{l1}, ..., \gamma_{lp}]^T \in \{0,1\}^p$ is a binary vector with $\gamma_{lj} = 1$ indicating the $l$-th causal signal is attributed to the $j$-th SNP, $b_l$ is the effect size of of the $l$-th causal signal, and $\boldsymbol{\pi}_l = [\pi_{l1}, ..., \pi_{lp}]^T \in [0,1]^p$ is the vector of prior causal probabilities with $\sum_{j=1}^{p} \pi_{lj} = 1$.

Consider the matrix of functional annotations $\mathbf{A} = [\mathbf{A}_1, ..., \mathbf{A}_p]^T \in \mathbb{R}^{p \times m}$ that collects $m$ annotations of the $p$ SNPs. We link the prior probability $\boldsymbol{\pi}_l$ to SNP annotations with the following softmax model with random effects:

$$\pi_{lj} = \frac{e^{\mathbf{A}_j^T \mathbf{w}_l}}{\sum_{j'=1}^{p} e^{\mathbf{A}_{j'}^T \mathbf{w}_l}}, \quad \mathbf{w}_l \sim \mathcal{N}(0, \sigma_{\mathbf{w}l}^2 \mathbf{I}_m), \tag{3}$$

where $\mathbf{w}_l \in \mathbb{R}^m$ represents the random effects vector of annotations on the causal probability of the $l$-th single-effect component.

We denote the set of model parameters $\boldsymbol{\theta} := \{\sigma^2, \boldsymbol{\sigma}_b^2, \boldsymbol{\sigma}_w^2\}$, where $\boldsymbol{\sigma}^2 := \{\sigma_{b1}^2, \cdots, \sigma_{bL}^2\}$ and $\boldsymbol{\sigma}_w^2 := \{\sigma_{w1}^2, \cdots, \sigma_{wL}^2\}$. The collections of random variables are denoted as $\tilde{\mathbf{b}} := \{b_1, \cdots, b_L\}$, $\tilde{\boldsymbol{\gamma}} := \{\boldsymbol{\gamma}_1, \cdots, \boldsymbol{\gamma}_L\}$, $\tilde{\mathbf{w}} := \{\mathbf{w}_1, \cdots, \mathbf{w}_L\}$. The logarithm of the marginal likelihood is given as:

$$\log \Pr(\mathbf{y} \mid \mathbf{X}, \mathbf{A}; \boldsymbol{\theta}) = \log \sum_{\tilde{\boldsymbol{\gamma}}} \int_{\tilde{\mathbf{b}}} \int_{\tilde{\mathbf{w}}} \Pr(\mathbf{y}, \tilde{\mathbf{b}}, \tilde{\boldsymbol{\gamma}}, \tilde{\mathbf{w}} \mid \mathbf{X}, \mathbf{A}; \boldsymbol{\theta}) d\tilde{\mathbf{w}} d\tilde{\mathbf{b}} \tag{4}$$

By maximizing the log-likelihood, we aim to obtain parameter estimates $\hat{\boldsymbol{\theta}}$ and prioritize the causal SNPs by using the posterior probability:

$$\Pr(\tilde{\boldsymbol{\gamma}}, \tilde{\mathbf{b}}, \tilde{\mathbf{w}} \mid \mathbf{y}, \mathbf{X}, \mathbf{A}; \hat{\boldsymbol{\theta}}) = \frac{\Pr(\tilde{\boldsymbol{\gamma}}, \tilde{\mathbf{b}}, \tilde{\mathbf{w}}, \mathbf{y} \mid \mathbf{X}, \mathbf{A}; \hat{\boldsymbol{\theta}})}{\Pr(\mathbf{y} \mid \mathbf{X}, \mathbf{A}; \hat{\boldsymbol{\theta}})}. \tag{5}$$

#### 1.2 Variational lower bound and algorithm

The marginal likelihood is intractable due the softmax function and sum-of-single-efffects assumption. To estimate model parameters $\boldsymbol{\theta}$ and approximate the posterior (5), we first derive a tractable lower bound of the log-likelihood. We show that an efficient algorithm can be developed based on the variational lower bound.

##### Derivation of Funmap variational lower bound

The marginal likelihood of Funmap model is given as

$$\begin{aligned}\Pr(\mathbf{y} \mid \mathbf{X}, \mathbf{A}; \boldsymbol{\theta}) &= \sum_{\tilde{\boldsymbol{\gamma}}} \int_{\tilde{\mathbf{b}}} \int_{\tilde{\mathbf{w}}} \Pr(\mathbf{y}, \tilde{\mathbf{b}}, \tilde{\boldsymbol{\gamma}}, \tilde{\mathbf{w}} \mid \mathbf{X}, \mathbf{A}; \boldsymbol{\theta}) d\tilde{\mathbf{w}} d\tilde{\mathbf{b}} \\ &= \sum_{\tilde{\boldsymbol{\gamma}}} \int_{\tilde{\mathbf{b}}} \int_{\tilde{\mathbf{w}}} \Pr(\mathbf{y} \mid \tilde{\mathbf{b}}, \tilde{\boldsymbol{\gamma}}, \tilde{\mathbf{w}}, \mathbf{X}; \boldsymbol{\theta}) \Pr(\tilde{\mathbf{b}}, \tilde{\boldsymbol{\gamma}} \mid \tilde{\mathbf{w}}, \mathbf{A}; \boldsymbol{\theta}) \Pr(\tilde{\mathbf{w}} \mid \boldsymbol{\theta}) d\tilde{\mathbf{w}} d\tilde{\mathbf{b}},\end{aligned} \tag{6}$$

where

$$\begin{aligned}\Pr(\mathbf{y} \mid \tilde{\mathbf{b}}, \tilde{\boldsymbol{\gamma}}, \tilde{\mathbf{w}}, \mathbf{X}; \boldsymbol{\theta}) &= (2\pi\sigma^2)^{-\frac{n}{2}} \exp(-\frac{1}{2\sigma^2}(\mathbf{y} - \mathbf{X}\mathbf{b})^T(\mathbf{y} - \mathbf{X}\mathbf{b})) \\ \Pr(\tilde{\mathbf{b}}, \tilde{\boldsymbol{\gamma}} \mid \tilde{\mathbf{w}}, \mathbf{A}; \boldsymbol{\theta}) &= \prod_{l=1}^{L} \frac{1}{\sqrt{2\pi\sigma_{bl}^2}} \exp(\frac{-b_l^2}{2\sigma_{bl}^2}) \cdot \prod_{j=1}^{p} \left[\frac{e^{\mathbf{A}_j^T \mathbf{w}_l}}{\sum_{j'=1}^{p} e^{\mathbf{A}_{j'}^T \mathbf{w}_l}}\right]^{\gamma_{lj}} \\ \Pr(\tilde{\mathbf{w}} \mid \boldsymbol{\theta}) &= \prod_{l=1}^{L} (2\pi\sigma_{\mathbf{w}l}^2)^{-\frac{m}{2}} \exp(-\frac{1}{2\sigma_{\mathbf{w}l}^2} \mathbf{w}_l^T \mathbf{w}_l).\end{aligned} \tag{7}$$

Due to the sum-of-exponential term on the denominator of softmax function, the integration over softmax function is intractable. We consider the double majorization of sum-of-exponential function [2]:

$$\sum_{j'=1}^{p} e^{\mathbf{A}_{j'}^T \mathbf{w}_l} \leq \exp[\rho_l + \sum_{j'=1}^{p} \frac{\mathbf{A}_{j'}^T \mathbf{w}_l - \rho_l - \xi_{lj'}}{2} + \lambda(\xi_{lj'})((\mathbf{A}_{j'}^T \mathbf{w}_l - \rho_l)^2 - \xi_{lj'}^2) + \log(1 + e^{\xi_{lj'}})], \tag{8}$$

where $\lambda(\xi_{lj'}) = \frac{1}{2\xi_{lj'}}(\frac{1}{1+e^{-\xi_{lj'}}} - \frac{1}{2})$, $\xi_{lj'} \in [0, \infty)$, and $\rho \in \mathbb{R}$. Clearly, the right-hand side of this inequality is in the exponentiated quadratic form. Therefore, applying this inequality to the denominator of the softmax function (3) leads to a tractable bound

$$\begin{aligned}&\Pr(\tilde{\mathbf{b}}, \tilde{\boldsymbol{\gamma}} \mid \mathbf{A}, \tilde{\mathbf{w}}; \boldsymbol{\sigma}_b^2) \\ =& \prod_{l=1}^{L} \frac{1}{\sqrt{2\pi\sigma_{bl}^2}} \exp(\frac{-b_l^2}{2\sigma_{bl}^2}) \cdot \prod_{j=1}^{p} \left[\frac{e^{\mathbf{A}_j^T \mathbf{w}_l}}{\sum_{j'=1}^{p} e^{\mathbf{A}_{j'}^T \mathbf{w}_l}}\right]^{\gamma_{lj}} \\ \geq& \prod_{l=1}^{L} \frac{1}{\sqrt{2\pi\sigma_{bl}^2}} \exp(\frac{-b_l^2}{2\sigma_{bl}^2}) \cdot \\ &\prod_{j=1}^{p} \left(\frac{\exp(\mathbf{A}_j^T \mathbf{w}_l)}{\exp(\rho_l + \sum_{j'=1}^{p} [\frac{\mathbf{A}_{j'}^T \mathbf{w}_l - \rho_l - \xi_{lj'}}{2} + \lambda(\xi_{lj'})((\mathbf{A}_{j'}^T \mathbf{w}_l - \rho_l)^2 - \xi_{lj'}^2) + \log(1 + e^{\xi_{lj'}})])}\right)^{\gamma_{lj}} \\ \equiv& g(\tilde{\mathbf{b}}, \tilde{\boldsymbol{\gamma}} \mid \mathbf{A}, \tilde{\mathbf{w}}; \boldsymbol{\xi}, \boldsymbol{\rho}, \boldsymbol{\sigma}_b^2)\end{aligned} \tag{9}$$

where $\boldsymbol{\xi} \in [0, \infty)^{L \times p}$ and $\boldsymbol{\rho} \in \mathbb{R}^L$ are variational parameters. Let $\boldsymbol{\Theta} = \{\boldsymbol{\theta}, \boldsymbol{\xi}, \boldsymbol{\rho}\}$ collects the model parameters and variational parameters. We can obtain a lower bound of complete-data likelihood as:

$$
\begin{aligned}
\Pr(\mathbf{y}, \tilde{\mathbf{b}}, \tilde{\boldsymbol{\gamma}}, \tilde{\mathbf{w}} \mid \mathbf{X}, \mathbf{A}; \boldsymbol{\theta}) &= \Pr(\mathbf{y} \mid \mathbf{X}, \tilde{\mathbf{b}}, \tilde{\boldsymbol{\gamma}}, \tilde{\mathbf{w}}; \sigma^2) \cdot \Pr(\tilde{\mathbf{b}}, \tilde{\boldsymbol{\gamma}} \mid \mathbf{A}, \tilde{\mathbf{w}}; \boldsymbol{\sigma}_b^2) \cdot \Pr(\tilde{\mathbf{w}}; \boldsymbol{\sigma}_w^2) \\
&\geq \Pr(\mathbf{y} \mid \mathbf{X}, \tilde{\mathbf{b}}, \tilde{\boldsymbol{\gamma}}, \tilde{\mathbf{w}}; \sigma^2) \cdot g(\tilde{\mathbf{b}}, \tilde{\boldsymbol{\gamma}} \mid \mathbf{A}, \tilde{\mathbf{w}}; \boldsymbol{\xi}, \boldsymbol{\rho}, \boldsymbol{\sigma}_b^2) \cdot \Pr(\tilde{\mathbf{w}}; \boldsymbol{\sigma}_w^2) \\
&\equiv f(\mathbf{y}, \tilde{\mathbf{b}}, \tilde{\boldsymbol{\gamma}}, \tilde{\mathbf{w}} \mid \mathbf{X}, \mathbf{A}; \boldsymbol{\Theta})
\end{aligned} \tag{10}
$$

Based on the above bound, we further derive a lower bound of the logarithm of marginal likelihood

$$
\begin{aligned}
&\log \Pr(\mathbf{y} \mid \mathbf{X}, \mathbf{A}; \boldsymbol{\theta}) \\
=&\log \sum_{\tilde{\boldsymbol{\gamma}}} \int_{\tilde{\mathbf{b}}} \int_{\tilde{\mathbf{w}}} \Pr(\mathbf{y}, \tilde{\mathbf{b}}, \tilde{\boldsymbol{\gamma}}, \tilde{\mathbf{w}} \mid \mathbf{X}, \mathbf{A}; \boldsymbol{\theta}) d\tilde{\mathbf{w}}\ d\tilde{\mathbf{b}} \\
\geq&\log \sum_{\tilde{\boldsymbol{\gamma}}} \int_{\tilde{\mathbf{b}}} \int_{\tilde{\mathbf{w}}} f(\mathbf{y}, \tilde{\mathbf{b}}, \tilde{\boldsymbol{\gamma}}, \tilde{\mathbf{w}} \mid \mathbf{X}, \mathbf{A}; \boldsymbol{\Theta}) d\tilde{\mathbf{w}}\ d\tilde{\mathbf{b}} \\
\geq&\sum_{\tilde{\boldsymbol{\gamma}}} \int_{\tilde{\mathbf{b}}} \int_{\tilde{\mathbf{w}}} q(\tilde{\mathbf{b}}, \tilde{\boldsymbol{\gamma}}, \tilde{\mathbf{w}}) \log \frac{f(\mathbf{y}, \tilde{\mathbf{b}}, \tilde{\boldsymbol{\gamma}}, \tilde{\mathbf{w}} \mid \mathbf{X}, \mathbf{A}; \boldsymbol{\Theta})}{q(\tilde{\mathbf{b}}, \tilde{\boldsymbol{\gamma}}, \tilde{\mathbf{w}})} d\tilde{\mathbf{w}}\ d\tilde{\mathbf{b}} \\
=&\mathbb{E}_q[\log f(\boldsymbol{y}, \tilde{b}, \tilde{\boldsymbol{\gamma}}, \tilde{\boldsymbol{w}} \mid \mathbf{X}, \mathbf{A}; \boldsymbol{\Theta}) - \log q(\tilde{\mathbf{b}}, \tilde{\boldsymbol{\gamma}}, \tilde{\mathbf{w}})] \\
\equiv&F(q, \boldsymbol{\Theta}),
\end{aligned} \tag{11}
$$

where the first inequality follows the double majorization bound, the second inequality is granted by Jensen's inequality, and $q(\tilde{\mathbf{b}}, \tilde{\boldsymbol{\gamma}}, \tilde{\mathbf{w}})$ is an approximation of the exact posterior (5). To analytically evaluate the lower bound, we introduce the following factorizable mean-field formulation assumption:

$$
q(\tilde{\mathbf{b}}, \tilde{\boldsymbol{\gamma}}, \tilde{\mathbf{w}}) = \prod_{l=1}^{L} q_l(b_l, \boldsymbol{\gamma}_l, \mathbf{w}_l) = \prod_{l=1}^{L} q_l(b_l, \boldsymbol{\gamma}_l) q_l(\mathbf{w}_l) \tag{12}
$$

where $q_l(b_l, \boldsymbol{\gamma}_l) = q(b_l \mid \boldsymbol{\gamma}_l) q(\boldsymbol{\gamma}_l)$. The above approximation can be evaluated in closed form, which further allows the lower bound to be analytically evaluated. Besides, by taking advantage of the sum-of-single-effects decomposition, this mean-field assumption inherits the property of SuSiE that only requires the $L$ causal signals to be independent with each other, relaxing the assumptions in traditional variational approximations [3, 4]. Under assumption (12), the lower bound can be expressed as $F(q, \boldsymbol{\Theta}) = F(q_1, ..., q_L, \boldsymbol{\Theta})$. In the following, we develop a variational algorithm to maximize $F(q_1, ..., q_L, \boldsymbol{\Theta})$. Briefly, we show that $F(q_1, ..., q_L, \boldsymbol{\Theta})$ can be maximized by an iterative procedure that alternatingly updates each posterior $q_1, ..., q_L$ and model parameters $\boldsymbol{\theta}$ in turns until convergence.

##### Update of $q_l(b_l, \boldsymbol{\gamma}_l, \mathbf{w}_l)$

We first focus on the update of the $l$-th approximate posterior $q_l(b_l, \boldsymbol{\gamma}_l, \mathbf{w}_l)$. The optimal $q_l$ is given by the solution of the following optimization problem:

$$
\max_{q_l} F(q_1, \ldots, q_L, \boldsymbol{\Theta}). \tag{13}
$$

To simplify the problem, we note that the lower bound $F(q_1, ..., q_L, \mathbf{\Theta})$ can be analytically expressed as:

$$F(q_1, \ldots, q_L, \mathbf{\Theta}) = -\frac{n}{2}\log(2\pi\sigma^2) - \frac{1}{2\sigma^2}\mathbb{E}_q[\|\mathbf{y} - \sum_{l=1}^{L}\mathbf{X}\mathbf{b}_l\|^2]$$
$$+ \sum_{l=1}^{L}(\mathbb{E}_{q_l}[\log g(b_l, \boldsymbol{\gamma}_l \mid \mathbf{A}, \mathbf{w}_l; \xi_l, \rho_l, \sigma_{bl}^2) + \log \Pr(\mathbf{w}_l; \sigma_{wl}^2) - \log q_l(b_l, \boldsymbol{\gamma}_l, \mathbf{w}_l)]) \tag{14}$$

where

$$\log g(b_l, \boldsymbol{\gamma}_l \mid \mathbf{A}, \mathbf{w}_l; \xi_l, \rho_l, \sigma_{bl}^2) = -\frac{1}{2}\log(2\pi\sigma_{bl}^2) - \frac{1}{2\sigma_{bl}^2}b_l^2 + \sum_{j=1}^{p}\gamma_{lj}\mathbf{A}_j^T\mathbf{w}_l$$
$$- \sum_{j=1}^{p}\gamma_{lj}\{\rho_l + \sum_{j'=1}^{p}\frac{\mathbf{A}_{j'}^T\mathbf{w}_l - \rho_l - \xi_{lj'}}{2} + \sum_{j'=1}^{p}\lambda(\xi_{lj'})((\mathbf{A}_{j'}^T\mathbf{w}_l - \rho_l)^2 - \xi_{lj'}^2)$$
$$+ \sum_{j'=1}^{p}\log(1 + e^{\xi_{lj'}})\}$$
$$\log \Pr(\mathbf{w}_l; \sigma_{wl}^2) = -\frac{m}{2}\log(2\pi\sigma_{wl}^2) - \frac{1}{2\sigma_{wl}^2}\mathbf{w}_l^T\mathbf{w}_l.$$

We extract the terms involving $q_l$ by further decomposing Equation (14) into terms with and without $q_l$:

$$F(q_1, \ldots, q_L, \mathbf{\Theta}) = -\frac{n}{2}\log(2\pi\sigma^2) - \frac{1}{2\sigma^2}\mathbb{E}_q[\|\mathbf{y} - \sum_{l=1}^{L}\mathbf{X}\mathbf{b}_l\|^2]$$
$$+ \sum_{l=1}^{L}\mathbb{E}_{q_l}[\log g(b_l, \boldsymbol{\gamma}_l \mid \mathbf{A}, \mathbf{w}_l; \xi_l, \rho_l, \sigma_{bl}^2) + \log \Pr(\mathbf{w}_l; \sigma_{wl}^2) - \log q_l(b_l, \boldsymbol{\gamma}_l, \mathbf{w}_l)]$$
$$= -\frac{n}{2}\log(2\pi\sigma^2) - \frac{1}{2\sigma^2}\|\mathbf{y} - \sum_{l=1}^{L}\mathbf{X}\mathbb{E}_q[\mathbf{b}_l]\|^2 + \sum_{l=1}^{L}\mathrm{Var}_{q_l}(\mathbf{X}\mathbf{b}_l)$$
$$+ \sum_{l=1}^{L}\mathbb{E}_{q_l}[\log g(b_l, \boldsymbol{\gamma}_l \mid \mathbf{A}, \mathbf{w}_l; \xi_l, \rho_l, \sigma_{bl}^2) + \log \Pr(\mathbf{w}_l; \sigma_{wl}^2) - \log q_l(b_l, \boldsymbol{\gamma}_l, \mathbf{w}_l)]$$
$$= -\frac{n}{2}\log(2\pi\sigma^2) - \frac{1}{2\sigma^2}\|\mathbf{y} - \sum_{l'\neq l}^{L}\mathbf{X}\mathbb{E}_{q_{l'}}[\mathbf{b}_{l'}] - \mathbf{X}\mathbb{E}_q[\mathbf{b}_l]\|^2 + \sum_{l=1}^{L}\mathrm{Var}_{q_l}(\mathbf{X}\mathbf{b}_l)$$
$$+ \sum_{l=1}^{L}\mathbb{E}_{q_l}[\log g(b_l, \boldsymbol{\gamma}_l \mid \mathbf{A}, \mathbf{w}_l; \xi_l, \rho_l, \sigma_{bl}^2) + \log \Pr(\mathbf{w}_l; \sigma_{wl}^2) - \log q_l(b_l, \boldsymbol{\gamma}_l, \mathbf{w}_l)]$$
$$= -\frac{n}{2}\log(2\pi\sigma^2) - \frac{1}{2\sigma^2}\mathbb{E}_{q_l}[\|\bar{\boldsymbol{r}}_l - \mathbf{X}\mathbf{b}_l\|^2]$$
$$+ \mathbb{E}_{q_l}[\log g(b_l, \boldsymbol{\gamma}_l \mid \mathbf{A}, \mathbf{w}_l; \xi_l, \rho_l, \sigma_{bl}^2) + \log \Pr(\mathbf{w}_l; \sigma_{wl}^2) - \log q_l(b_l, \boldsymbol{\gamma}_l, \mathbf{w}_l)] + \text{const}$$
$$= F_l(q_l, \mathbf{\Theta}) + \text{const}, \tag{15}$$

where $\bar{\boldsymbol{r}}_l = \mathbf{y} - \sum_{l'\neq l}\mathbf{X}\mathbb{E}_{q_{l'}}[\mathbf{b}_{l'}]$ and

$$F_l(q_l, \mathbf{\Theta}) = -\frac{n}{2}\log(2\pi\sigma^2) - \frac{1}{2\sigma^2}\mathbb{E}_{q_l}[\|\bar{\boldsymbol{r}}_l - \mathbf{X}\mathbf{b}_l\|^2]$$
$$+ \mathbb{E}_{q_l}[\log g(b_l, \boldsymbol{\gamma}_l \mid \mathbf{A}, \mathbf{w}_l; \xi_l, \rho_l, \sigma_{bl}^2) + \log \Pr(\mathbf{w}_l; \sigma_{wl}^2) - \log q_l(b_l, \boldsymbol{\gamma}_l, \mathbf{w}_l)],$$

According to Equation (15), optimizing the lower bound $F(q_1, \dots, q_L, \boldsymbol{\Theta})$ with respect to $q_l$ is equivalent to optimizing $F_l(q_l, \boldsymbol{\Theta})$ with other $q_{l'}$ fixed:

$$\underset{q_l}{\operatorname{argmax}} \ F(q_1, \dots, q_L, \boldsymbol{\Theta}) = \underset{q_l}{\operatorname{argmax}} \ F_l(q_l, \boldsymbol{\Theta}). \tag{16}$$

With the above equivalence, we can sequentially update $q_l$ by working on $F_l(q_l, \boldsymbol{\Theta})$ in turns.

Now, we show that the above problem is equivalent to fitting a funcationally informed single-effect regression (FunSER) with $\mathbf{y}$ replaced by $\bar{\boldsymbol{r}}_l$. Specifically, this FunSER aims to maximize the following likelihood:

$$\begin{aligned} \Pr(\bar{\boldsymbol{r}} \mid \mathbf{X}, \mathbf{A}; \boldsymbol{\theta}) &= \sum_{\boldsymbol{\gamma}_l} \int_{b_l} \int_{\mathbf{w}_l} \Pr(\bar{\boldsymbol{r}}, b_l, \boldsymbol{\gamma}_l, \mathbf{w}_l \mid \mathbf{X}, \mathbf{A}; \boldsymbol{\theta}) d\mathbf{w}_l db_l \\ =& \sum_{\boldsymbol{\gamma}_l} \int_{b_l} \int_{\mathbf{w}_l} \Pr(\bar{\boldsymbol{r}} \mid b_l, \boldsymbol{\gamma}_l, \mathbf{w}_l, \mathbf{X}; \boldsymbol{\theta}) \Pr(b_l, \boldsymbol{\gamma}_l \mid \mathbf{w}_l, \mathbf{A}; \boldsymbol{\theta}) \Pr(\mathbf{w}_l \mid \boldsymbol{\theta}) d\mathbf{w}_l db_l, \end{aligned} \tag{17}$$

where

$$\begin{aligned} &\Pr(\bar{\boldsymbol{r}} \mid b_l, \boldsymbol{\gamma}_l, \mathbf{w}_l, \mathbf{X}; \boldsymbol{\theta}) = (2\pi\sigma^2)^{-\frac{n}{2}} \exp(-\frac{1}{2\sigma^2}(\bar{\boldsymbol{r}} - \mathbf{X} b_l \boldsymbol{\gamma}_l)^T (\bar{\boldsymbol{r}} - \mathbf{X} b_l \boldsymbol{\gamma}_l)) \\ &\Pr(b_l, \boldsymbol{\gamma}_l \mid \mathbf{w}_l, \mathbf{A}; \boldsymbol{\theta}) = \frac{1}{\sqrt{2\pi\sigma_{bl}^2}} \exp(\frac{-b_l^2}{2\sigma_{bl}^2}) \cdot \prod_{j=1}^{p} \left[ \frac{e^{\mathbf{A}_j^T \mathbf{w}_l}}{\sum_{j'=1}^{p} e^{\mathbf{A}_{j'}^T \mathbf{w}_l}} \right]^{\gamma_{lj}} \\ &\Pr(\mathbf{w}_l \mid \boldsymbol{\theta}) = (2\pi\sigma_{\mathbf{w}l}^2)^{-\frac{m}{2}} \exp(-\frac{1}{2\sigma_{\mathbf{w}l}^2} \mathbf{w}_l^T \mathbf{w}_l). \end{aligned} \tag{18}$$

By applying the double majorization bound (8) as we did in the full Funmap model, we can obtain a lower bound of the complete-data likelihood for FunSER

$$\begin{aligned} &\log \Pr(\bar{\boldsymbol{r}}, b_l, \boldsymbol{\gamma}_l, \mathbf{w}_l \mid \mathbf{X}, \mathbf{A}; \boldsymbol{\theta}) \\ =& \log \Pr(\bar{\boldsymbol{r}} \mid \boldsymbol{X}, b_l, \boldsymbol{\gamma}_l, \mathbf{w}_l; \sigma^2) + \log \Pr(b_l, \boldsymbol{\gamma}_l \mid \mathbf{A}, \mathbf{w}_l; \sigma_{bl}^2) + \log \Pr(\mathbf{w}_l \mid \sigma_{wl}^2) \\ \geq& \log \Pr(\bar{\boldsymbol{r}} \mid \mathbf{X}, b_l, \boldsymbol{\gamma}_l, \mathbf{w}_l; \sigma^2) + \log g(b_l, \boldsymbol{\gamma}_l \mid \mathbf{A}, \mathbf{w}_l; \xi_l, \rho_l, \sigma_{bl}^2) + \log \Pr(\mathbf{w}_l \mid \sigma_{wl}^2) \\ \equiv& \log f(\bar{\boldsymbol{r}}, b_l, \boldsymbol{\gamma}_l, \mathbf{w}_l \mid \mathbf{X}, \mathbf{A}; \boldsymbol{\Theta}). \end{aligned} \tag{19}$$

Combining the above bound and the Jensen's inequality, the lower bound of the logarithm of FunSER likelihood (17) can be obtained as

$$\begin{aligned} &\log \Pr(\bar{\boldsymbol{r}} \mid \mathbf{X}, \mathbf{A}; \boldsymbol{\theta}) \\ \geq& \log \sum_{\boldsymbol{\gamma}_l} \int_{b_l} \int_{\mathbf{w}_l} f(\bar{\boldsymbol{r}}, b_l, \boldsymbol{\gamma}_l, \mathbf{w}_l \mid \mathbf{X}, \mathbf{A}; \boldsymbol{\Theta}) d\mathbf{w}_l \ db_l \\ \geq& \sum_{\boldsymbol{\gamma}_l} \int_{b_l} \int_{\mathbf{w}_l} q(b_l, \boldsymbol{\gamma}_l, \mathbf{w}_l) \log \frac{f(\bar{\boldsymbol{r}}, b_l, \boldsymbol{\gamma}_l, \mathbf{w}_l \mid \mathbf{X}, \mathbf{A}; \boldsymbol{\Theta})}{q_l(b_l, \boldsymbol{\gamma}_l, \mathbf{w}_l)} d\mathbf{w}_l \ db_l \\ =& \mathbb{E}_{q_l}[\log f(\bar{\boldsymbol{r}}, b_l, \boldsymbol{\gamma}_l, \mathbf{w}_l \mid \mathbf{X}, \mathbf{A}; \boldsymbol{\Theta}) - \log q_l(b_l, \boldsymbol{\gamma}_l, \mathbf{w}_l)] \\ =& F_l(q_l, \boldsymbol{\Theta}), \end{aligned} \tag{20}$$

As we can see, this FunSER lower bound is equivalent to the $F_l(q_l, \boldsymbol{\Theta})$ defined in Equation(15). By inheriting the factorizable assumption (12) from full Funmap model, we have

$$q_l(b_l, \boldsymbol{\gamma}_l, \mathbf{w}_l) = q_l(b_l, \boldsymbol{\gamma}_l) q_l(\mathbf{w}_l) \tag{21}$$

where $q_l(b_l, \boldsymbol{\gamma}_l) = q(b_l \mid \boldsymbol{\gamma}_l)q(\boldsymbol{\gamma}_l)$ and $q(\gamma_{lj} = 1) = \alpha_{lj}$. Based on the assumption (21) and lower bound (20), we can obtain a closed form expression of $\log q_l(b_l, \boldsymbol{\gamma}_l)$:

$$
\begin{aligned}
\log q_l(b_l, \boldsymbol{\gamma}_l) &= \mathbb{E}_{\mathbf{w}_l}\left[\log f(\bar{\boldsymbol{r}}_l, b_l, \boldsymbol{\gamma}_l, \mathbf{w}_l \mid \mathbf{X}, \mathbf{A}; \boldsymbol{\theta})\right] \\
&= -\frac{1}{2\sigma^2}(-2b_l\boldsymbol{\gamma}_l^T\mathbf{X}^T\bar{\boldsymbol{r}}_l + b_l\boldsymbol{\gamma}_l^T\mathbf{X}^T\mathbf{X}b_l\boldsymbol{\gamma}_l) - \frac{1}{2\sigma_{bl}^2}b_l^2 + \text{const} \\
&= (-\frac{1}{2\sigma^2}\boldsymbol{\gamma}_l^T\mathbf{X}^T\mathbf{X}\boldsymbol{\gamma}_l - \frac{1}{2\sigma_{bl}^2})b_l^2 + \frac{1}{\sigma^2}\boldsymbol{\gamma}_l^T\mathbf{X}^T\bar{\boldsymbol{r}}_l b_l,
\end{aligned} \tag{22}
$$

where the expectation is taken under the distribution $q_l(\mathbf{w}_l)$. When $\gamma_{lj} = 1$, we have

$$
\log q_l(b_l \mid \gamma_{lj} = 1) = (-\frac{1}{2\sigma^2}\mathbf{x}_j^T\mathbf{x}_j - \frac{1}{2\sigma_{bl}^2})b_l^2 + \frac{1}{\sigma^2}\mathbf{x}_j^T\bar{\boldsymbol{r}}_l b_l, \tag{23}
$$

which is a quadratic form. Thus, we can obtain the posterior $q_l(b_l, \boldsymbol{\gamma}_l)$ as

$$
q_l(b_l, \boldsymbol{\gamma}_l) \sim \prod_{j=1}^{p}(\alpha_{lj}\mathcal{N}(\mu_{blj}, s_{blj}^2))^{\gamma_{lj}} \tag{24}
$$

where

$$
s_{blj}^2 = \frac{1}{\frac{1}{\sigma^2}\mathbf{x}_j^T\mathbf{x}_j + \frac{1}{\sigma_{b_l}^2}}, \ \mu_{blj} = s_{blj}^2 \cdot \frac{1}{\sigma^2}\mathbf{x}_j^T\bar{\boldsymbol{r}}_l.
$$

Similarly, we can obtain a closed form expression of $\log q_l\left(\mathbf{w}_l\right)$:

$$
\begin{aligned}
\log q_l\left(\mathbf{w}_l\right) &= \mathbb{E}_{b_l, \boldsymbol{\gamma}_l}\left[\log f(\bar{\boldsymbol{r}}_l, b_l, \boldsymbol{\gamma}_l, \boldsymbol{w}_l \mid \mathbf{X}, \mathbf{A}; \boldsymbol{\theta})\right] \\
&= \mathbf{w}_l^T\left(-\frac{1}{2\sigma_{wl}^2}\mathbf{I}_M - \sum_{j=1}^{p}\lambda\left(\xi_{lj}\right)\mathbf{A}_j\mathbf{A}_j^T\right)\mathbf{w}_l \\
&+ \mathbf{w}_l^T\sum_{j=1}^{p}\left(\alpha_{lj} - \frac{1}{2} + 2\rho_l\lambda\left(\xi_{lj}\right)\right)\mathbf{A}_j \\
&+ \text{const},
\end{aligned} \tag{25}
$$

where the expectation is taken over the distribution $q_l(b_l, \boldsymbol{\gamma}_l)$. Again, the above quadratic form indicates that

$$
q_l\left(\mathbf{w}_l\right) \sim \mathcal{N}_m(\boldsymbol{\mu}_{wl}, \boldsymbol{\Sigma}_{wl}), \tag{26}
$$

where

$$
\boldsymbol{\Sigma}_{wl}^{-1} = \frac{1}{\sigma_{wl}^2}\mathbf{I}_M + 2\sum_{j=1}^{p}\lambda\left(\xi_{lj}\right)\mathbf{A}_j\mathbf{A}_j^T, \ \boldsymbol{\mu}_{wl} = \boldsymbol{\Sigma}_{wl}\sum_{j=1}^{p}\left(\alpha_{lj} - \frac{1}{2} + 2\rho_l\lambda\left(\xi_{lj}\right)\right)\mathbf{A}_j.
$$

With the posterior expressions (24) and (26), we can evaluate the $F_l(q_l, \boldsymbol{\Theta})$ as

$$
\begin{aligned}
F_l(q_l, \boldsymbol{\Theta}) &= \mathbb{E}_{q_l}[\log f(\bar{\boldsymbol{r}}_l, b_l, \boldsymbol{\gamma}_l, \mathbf{w}_l \mid \mathbf{X}, \mathbf{A}; \boldsymbol{\Theta}) - \log q_l(b_l, \boldsymbol{\gamma}_l, \mathbf{w}_l)] \\
&= \mathbb{E}_{q_l}[\log \Pr(\bar{\boldsymbol{r}}_l \mid \mathbf{X}, b_l, \boldsymbol{\gamma}_l, \mathbf{w}_l; \sigma^2)] + \mathbb{E}_{q_l}[\log g(b_l, \boldsymbol{\gamma}_l \mid \mathbf{A}, \mathbf{w}_l; \xi_l, \rho_l, \sigma_{bl}^2)] \\
&+ \mathbb{E}_{q_l}[\log \Pr(\mathbf{w}_l; \sigma_{wl}^2)] - \mathbb{E}_{q_l}[\log q_l(b_l, \boldsymbol{\gamma}_l)] - \mathbb{E}_{q_l}[\log q_l(\mathbf{w}_l)] \\
&= -\frac{n}{2}\log(2\pi\sigma^2) - \frac{1}{2\sigma^2}\|\bar{\boldsymbol{r}}_l - \sum_{j=1}^{p} \mathbf{x}_j \mu_{blj}\alpha_{lj}\|^2 \\
&- \frac{1}{2\sigma^2}\sum_{i=1}^{n}\sum_{j=1}^{p} x_{ij}^2(\mu_{blj}^2 + s_{blj}^2)\alpha_{lj} + \frac{1}{2\sigma^2}\sum_{i=1}^{n}(\sum_{j=1}^{p} x_{ij}\mu_{blj}\alpha_{lj})^2 \\
&- \frac{1}{2}\log(2\pi\sigma_{bl}^2) - \frac{1}{2\sigma_{bl}^2}\sum_{j=1}^{p}(\mu_{blj}^2 + s_{blj}^2)\alpha_{lj} + \sum_{j=1}^{p}\alpha_{lj}\mathbf{A}_j^T\boldsymbol{\mu}_{wl} - \rho_l - \sum_{j=1}^{p}\lambda(\xi_{lj})\mathbf{A}_j^T\boldsymbol{\Sigma}_{wl}\mathbf{A}_j \\
&- \sum_{j=1}^{p}\left(\frac{\mathbf{A}_j^T\boldsymbol{\mu}_{wl} - \rho_l - \xi_{lj}}{2} + \lambda(\xi_{lj})((\mathbf{A}_j^T\boldsymbol{\mu}_{wl} - \rho_l)^2 - \xi_{lj}^2) + \log(1 + e^{\xi_{lj}})\right) \\
&- \frac{m}{2}\log(2\pi\sigma_{wl}^2) - \frac{1}{2\sigma_{wl}^2}\mathrm{Tr}(\boldsymbol{\mu}_{wl}\boldsymbol{\mu}_{wl}^T + \boldsymbol{\Sigma}_{wl}) \\
&+ \sum_{j=1}^{p}\frac{1}{2}\alpha_{lj}\log(2\pi e s_{blj}^2) - \sum_{j=1}^{p}\alpha_{lj}\log\alpha_{lj} + \frac{1}{2}\log((2\pi e)^m|\boldsymbol{\Sigma}_{wl}|).
\end{aligned}
\tag{27}
$$

To update $\alpha_{lj}$, we set the partial derivative of $F_l(q_l, \boldsymbol{\Theta})$ w.r.t to $\alpha_{lj}$ be 0, which gives us

$$
\alpha_{lj} = \frac{\exp(u_j)}{\sum_{j'=1}^{p}\exp(u_{j'})}, \text{where } u_j = \mathbf{A}_j^T\boldsymbol{\mu}_{wl} + \frac{\mu_{blj}^2}{2s_{blj}^2} + \frac{1}{2}\log s_{blj}^2 \tag{28}
$$

Based on *Bouchard, 2008*, we can update the variational parameters $\rho_l$, $\xi_{lj}^2$ as follows:

$$
\begin{aligned}
\xi_{lj}^2 &= \mathbf{A}_j^T(\boldsymbol{\Sigma}_{wl} + \boldsymbol{\mu}_{wl}\boldsymbol{\mu}_{wl}^T)\mathbf{A}_j + \rho_l^2 - 2\rho_l\mathbf{A}_j^T\boldsymbol{\mu}_{wl}, \\
\rho_l &= \frac{\frac{1}{2}(\frac{p}{2} - 1) + \sum_{j=1}^{p}\lambda(\xi_{lj})\boldsymbol{A}_j^T\boldsymbol{\mu}_{wl}}{\sum_{j=1}^{p}\lambda(\xi_{lj})}.
\end{aligned}
\tag{29}
$$

**Update of $\sigma_{bl}^2$ and $\sigma_{wl}^2$**

Now we derive the updates of $\sigma_{bl}^2$ and $\sigma_{wl}^2$. We set the partial derivative of $F_l(q_l, \boldsymbol{\Theta})$ w.r.t the parameters to be 0 and get:

$$
\sigma_{bl}^2 = \sum_{j=1}^{p}\alpha_{lj}(\mu_{blj}^2 + s_{blj}^2) \tag{30}
$$

$$
\sigma_{wl}^2 = \frac{\mathrm{Tr}(\boldsymbol{\Sigma}_{wl} + \boldsymbol{\mu}_{wl}\boldsymbol{\mu}_{wl}^T)}{m} \tag{31}
$$

**Update of $\sigma^2$**

The update for $\sigma^2$ is easily obtained by setting partial derivative of (14) with respect to $\sigma^2$ to zero

$$
\hat{\sigma}^2 = \frac{1}{n}\mathbb{E}_q[\|\mathbf{y} - \sum_{l=1}^{L}\mathbf{X}\mathbf{b}_l\|^2] \tag{32}
$$

The expected value of $\|\mathbf{y} - \sum_{l=1}^{L} \boldsymbol{X}\boldsymbol{b}_l\|^2$ is the expected residual sum of squares (ERSS) under the variational approximation $q$, and depends on $q$ only through its first and second moments. We can derive the expression of ERSS as follows:

$$
\begin{aligned}
\text{ERSS} &= \mathbb{E}_q[\|\mathbf{y} - \sum_{l=1}^{L} \mathbf{X}\mathbf{b}_l\|^2] \\
&= \mathbb{E}_q[\mathbf{y}^T\mathbf{y} - 2\boldsymbol{y}^T \sum_{l=1}^{L} \mathbf{X}\mathbf{b}_l + \sum_{l=1}^{L}\sum_{l'=1}^{L} (\mathbf{X}\mathbf{b}_l)^T(\mathbf{X}\mathbf{b}_{l'})] \\
&= \mathbf{y}^T\mathbf{y} - 2\mathbf{y}^T\mathbb{E}_q[\sum_{l=1}^{L} \mathbf{X}\mathbf{b}_l] + \mathbb{E}_q[\sum_{l=1}^{L}\sum_{l'=1}^{L} (\mathbf{X}\mathbf{b}_l)^T(\mathbf{X}\mathbf{b}_{l'})] \\
&= \mathbf{y}^T\mathbf{y} - 2\mathbf{y}^T \sum_{l=1}^{L} \mathbb{E}_{q_l}[\mathbf{X}\mathbf{b}_l] + \sum_{l=1}^{L}\sum_{l'=1}^{L} (\mathbb{E}_{q_l}[(\mathbf{X}\mathbf{b}_l)^T]\mathbb{E}_{q_{l'}}[\mathbf{X}\mathbf{b}_{l'}]) \\
&- \sum_{l=1}^{L} (\mathbb{E}_{q_l}[(\mathbf{X}\mathbf{b}_l)^T]\mathbb{E}_{q_l}[\mathbf{X}\mathbf{b}_l]) + \sum_{l=1}^{L} (\mathbb{E}_{q_l}[(\mathbf{X}\mathbf{b}_l)^T(\mathbf{X}\mathbf{b}_l)]) \\
&= \|\mathbf{y} - \sum_{l=1}^{L} \mathbb{E}_{q_l}[\mathbf{X}\mathbf{b}_l]\|^2 - \sum_{l=1}^{L} (\mathbb{E}_{q_l}[(\mathbf{X}\mathbf{b}_l)^T]\mathbb{E}_{q_l}[\mathbf{X}\mathbf{b}_l]) + \sum_{l=1}^{L} (\mathbb{E}_{q_l}[(\mathbf{X}\mathbf{b}_l)^T(\mathbf{X}\mathbf{b}_l)]) \\
&= \|\mathbf{y} - \sum_{l=1}^{L} \mathbb{E}_{q_l}[\mathbf{X}\mathbf{b}_l]\|^2 + \sum_{l=1}^{L}\sum_{i=1}^{n} \text{Var}_{q_l}[\mathbf{x}_i^T\mathbf{b}_l]
\end{aligned}
\tag{33}
$$

where

$$
\mathbb{E}_{q_l}[\mathbf{X}\mathbf{b}_l] = \sum_{j=1}^{p} \mathbf{x}_j \mathbb{E}_{q_l}[b_l\gamma_{lj}] = \sum_{j=1}^{p} \mathbf{x}_j \mu_{blj}\alpha_{lj}, \tag{34}
$$

$$
\begin{aligned}
\text{Var}_q[\mathbf{x}_i^T\mathbf{b}_l] &= \mathbb{E}_{q_l}[(\mathbf{x}_i^T\mathbf{b}_l)^2] - (\mathbb{E}_{q_l}[\mathbf{x}_i^T\boldsymbol{b}_l])^2 \\
&= \mathbb{E}_{q_l}[(\sum_{j=1}^{p} x_{ij}b_l\gamma_{lj})^2] - (\mathbb{E}_{q_l}[\sum_{j=1}^{p} x_{ij}b_l\gamma_{lj}])^2 \\
&= \mathbb{E}_{q_l}[(\sum_{j=1}^{p}(x_{ij}^2 b_l^2\gamma_{lj}^2) + 2\sum_{j=1}^{p}\sum_{j'=1}^{p} x_{ij}x_{ij'}b_l\gamma_{lj}b_l\gamma_{lj'}] - (\mathbb{E}_{q_l}[\sum_{j=1}^{p} x_{ij}b_l\gamma_{lj}])^2 \\
&= \mathbb{E}_{q_l}[(\sum_{j=1}^{p}(x_{ij}^2 b_l^2\gamma_{lj}^2)] - (\mathbb{E}_{q_l}[\sum_{j=1}^{p} x_{ij}b_l\gamma_{lj}])^2 \\
&= \sum_{j=1}^{p} x_{ij}^2\mathbb{E}_{q_l}[b_l^2\gamma_{lj}^2] - (\sum_{j=1}^{p} x_{ij}\mathbb{E}_{q_l}[b_l\gamma_{lj}])^2 \\
&= \sum_{j=1}^{p} x_{ij}^2\mathbb{E}_{q_l}[b_l^2\gamma_{lj}^2] - (\sum_{j=1}^{p} x_{ij}\mathbb{E}_{q_l}[b_l\gamma_{lj}])^2 \\
&= \sum_{j=1}^{p} x_{ij}^2(\mathbb{E}_{q_l}[b_l]^2 + \text{Var}_{q_l}[b_l])(\mathbb{E}_{q_l}[\gamma_{lj}]^2 + \text{Var}_{q_l}[\gamma_{lj}]) - (\sum_{j=1}^{p} x_{ij}\mathbb{E}_{q_l}[b_l]\mathbb{E}_{q_l}[\gamma_{lj}])^2 \\
&= \sum_{j=1}^{p} x_{ij}^2(\mu_{blj}^2 + s_{blj}^2)(\alpha_{lj}^2 + \alpha_{lj}(1-\alpha_{lj})) - (\sum_{j=1}^{p} x_{ij}\mu_{blj}\alpha_{lj})^2 \\
&= \sum_{j=1}^{p} x_{ij}^2(\mu_{1lj}^2 + s_{blj}^2)\alpha_{lj} - (\sum_{j=1}^{p} x_{ij}\mu_{blj}\alpha_{lj})^2
\end{aligned}
\tag{35}
$$

Combing the updates of $q_l(b_l, \boldsymbol{\gamma}_l, \mathbf{w}_l)$, $\sigma_{bl}^2$ and $\sigma_{wl}^2$, and $\sigma^2$ lead to the iterative Bayesian stepwise selection (IBSS) algorithm for Funmap.

#### 1.3 Adaptation to summary statistics

While the variational EM algorithm proposed above is for individual-level genotype $\mathbf{X}$ and phenotype $\mathbf{y}$, it can be easily extended to using only GWAS summary data as input. Let us consider the $z$-scores obtained from marginal regressions:

$$z_j = \frac{\hat{\beta}_j}{\hat{s}_j}, \text{ where } \hat{\beta}_j = (\mathbf{x}_j^T \mathbf{x}_j)^{-1} \mathbf{x}_j^T \mathbf{y}, \; \hat{s}_j = \sqrt{\frac{||\mathbf{y} - \mathbf{x}_j \hat{\beta}_j||_2^2}{n \mathbf{x}_j^T \mathbf{x}_j}}, \tag{36}$$

and $\mathbf{x}_j \in \mathbb{R}^p$ is the $j$-th column of $\mathbf{X}$. We first note that the likelihood and its lower bound (11) depend on the GWAS data $\{\mathbf{X}, \mathbf{y}\}$ only through the sufficient statistics $\mathbf{X}^T\mathbf{X}$, $\mathbf{X}^T\mathbf{y}$ and $\mathbf{y}^T\mathbf{y}$. Since both $\mathbf{X}$ and $\mathbf{y}$ have been standardized, we can replace the sufficient statistics with $z$-scores and LD matrix $\mathbf{R}$ [1] with the following relationships:

$$\mathbf{X}^T\mathbf{X} = n\mathbf{R}, \quad \mathbf{X}^T\mathbf{y} = \sqrt{n}\tilde{\mathbf{z}}, \quad \mathbf{y}^T\mathbf{y} = n, \tag{37}$$

where

$$\tilde{z}_j = \boldsymbol{D}_j^{1/2} z_j, \; \boldsymbol{D}_j = \frac{n}{n + z_j^2},$$

and $\mathbf{R}$ can be computed with genotypes from a subset of GWAS samples or from a reference panel of similar ancestry background. Note that $\boldsymbol{D}_j$ has the interpretation as being $1-$PVE (Proportion of phenotypic Variance Explained) [5]. Therefore, $\tilde{\mathbf{z}}$ is also known as the vector of the PVE-adjusted $z$-scores. If all the effects are small, the estimated PVEs will be close to zero and we have $\boldsymbol{D}_j \approx 1$ and $\tilde{\mathbf{z}} \approx \mathbf{z}$. In addition to above quantities, individual-level IBSS algorithm requires $\bar{\boldsymbol{r}}_l$ to evaluate $\mathbf{X}^T\bar{\boldsymbol{r}}_l$ to update $q_l(b_l, \boldsymbol{\gamma}_l)$ (Equation (24)), which is not feasible when only summary-level data is available. Therefore, instead of operating on the expected residuals $\bar{\boldsymbol{r}}_l$, we keep tracks on $\tilde{\boldsymbol{r}}_l$ defined as

$$\tilde{\boldsymbol{r}}_l = \mathbf{X}^T\bar{\boldsymbol{r}}_l = \mathbf{X}^T\mathbf{y} - \sum_{l' \neq l} \mathbf{X}^T\mathbf{X}\mathbb{E}_{q_{l'}}[\mathbf{b}_{l'}], \tag{38}$$

which can be computed from the summary-level data.

#### 2 Details of the variational inference algorithm

The variational algorithm solves for a non-convex problem, which has multiple local optimal solutions. To make our algorithm more stable, we design a three-stage algorithm based on warm starts. In the first stage, we fit a special case of Funmap without including the functional information, which is equivalent to SuSiE. We obtain the initial parameter estimates $\sigma^2$, $\{\sigma_{bl}^2\}_{l=1,\cdots,L}$, $\{\sigma_{wl}^2\}_{l=1,\cdots,L}$ and the initial posterior parameters $\{\mu_{blj}\}_{l=1,\cdots,L}^{j=1,\cdots,p}$, $\{s_{blj}^2\}_{l=1,\cdots,L}^{j=1,\cdots,p}$, $\{\alpha_{lj}\}_{l=1,\cdots,L}^{j=1,\cdots,p}$. Then, we use these estimated parameters as the initial values in the second stage to fit Funmap with $\alpha_{lj}$ fixed. Finally, we use the estimated parameters from the second stage as the initial values to fit the full Funmap model in the third stage. Details are as follows:

##### 2.1 Stage 1: SuSiE model

**Algorithm 1** IBSS algorithm for SuSiE model

**Input:** Data $\mathbf{X}^T\mathbf{X}$, $\mathbf{X}^T\mathbf{y}$, $\mathbf{y}^T\mathbf{y}$ (which can be calculated from $\mathbf{z}$, $\mathbf{R}$ and $n$)

Initialize: $\sigma^2 = 1$; $\{\sigma_{bl}^2\} = 0.2$; $\{\mu_{blj}\} = 0$; $\{\alpha_{lj}\} = 1/p$

**repeat**

$\tilde{\boldsymbol{r}} \leftarrow \mathbf{X}^T\mathbf{y} - \sum_{l=1}^{L} \mathbf{X}^T \sum_{j=1}^{p} \mathbf{x}_j \mu_{blj}\alpha_{lj}$

**for** $l = 1, \cdots, L$ **do**

$\tilde{\boldsymbol{r}}_l \leftarrow \tilde{\boldsymbol{r}} + \mathbf{X}^T \sum_{j=1}^{p} \mathbf{x}_j \mu_{blj}\alpha_{lj}$

$s_{blj}^2 \leftarrow 1/(\mathbf{X}_j^T\mathbf{X}_j/\sigma^2 + 1/\sigma_{bl}^2), j = 1, \cdots, p$

$\mu_{blj} \leftarrow s_{blj}^2 \cdot \tilde{\boldsymbol{r}}_{lj}/\sigma^2, j = 1, \cdots, p$, where $\tilde{\boldsymbol{r}}_{lj}$ is the $j$-th coordinate element of $\tilde{\boldsymbol{r}}_l$

$\alpha_{lj} \leftarrow \exp(u_j)/\sum_{j'=1}^{p} \exp(u_{j'})$, where $u_j = \dfrac{\mu_{blj}^2}{2s_{blj}^2} + \dfrac{1}{2}\log s_{blj}^2, j = 1, \cdots, p$

$\sigma_{bl}^2 \leftarrow \sum_{j=1}^{p} \alpha_{lj}(\mu_{blj}^2 + s_{blj}^2)$

$\tilde{\boldsymbol{r}} \leftarrow \tilde{\boldsymbol{r}}_l - \mathbf{X}^T \sum_{j=1}^{p} \mathbf{x}_j \mu_{blj}\alpha_{lj}$

**end for**

$\sigma^2 \leftarrow \text{ERSS}/n$, where ERSS is given by (35)

**until** convergence criterion satisfied

**Output:** $\sigma^2$, $\{\sigma_{bl}^2\}_{l=1,\cdots,L}$, $\{\mu_{blj}\}_{l=1,\cdots,L}^{j=1,\cdots,p}$, $\{s_{blj}^2\}_{l=1,\cdots,L}^{j=1,\cdots,p}$, $\{\alpha_{lj}\}_{l=1,\cdots,L}^{j=1,\cdots,p}$

##### 2.2 Stage 2: SuSiE model with random-effect

**Algorithm 2** IBSS algorithm for SuSiE model with random-effect

**Input:** Data $\mathbf{X}^T\mathbf{X}$, $\mathbf{X}^T\mathbf{y}$, $\mathbf{y}^T\mathbf{y}$ (which can be calculated from $\mathbf{z}$, $\mathbf{R}$ and $n$) and $\boldsymbol{A}$

**Input:** Initial value $\sigma^2$, $\{\sigma_{bl}^2\}$, $\{\mu_{blj}\}$, $\{s_{blj}^2\}$, $\{\alpha_{lj}\}$ from the Output of Stage 1

Initialize: $\{\boldsymbol{\mu}_{wl}\} = \mathbf{0}$; $\{\boldsymbol{\Sigma}_{wl}\} = \mathbf{I}_m$ and $\sigma^2$, $\{\sigma_{bl}^2\}$, $\{\mu_{blj}\}$, $\{s_{blj}^2\}$, $\{\alpha_{lj}\}$

Initialize: $\{\sigma_{wl}^2\}_{l=1,\cdots,l_s} = 0.1$; $\{\xi_{lj}\}_{l=1,\cdots,l_s} = (p/2-1)/2$; $\{\rho_l\}_{l=1,\cdots,l_s} = 0$

Initialize: $\{\sigma_{wl}^2\}_{l=l_s+1,\cdots,L} = 10^{-5}$; $\{\xi_{lj}\}_{l=l_s+1,\cdots,L} = 1$; $\{\rho_l\}_{l=l_s+1,\cdots,L} = 0$

where $l_s$ represents the number of credible sets determined by the SuSiE model.

**repeat**

$\tilde{\boldsymbol{r}} \leftarrow \mathbf{X}^T\boldsymbol{y} - \sum_{l=1}^{L} \mathbf{X}^T \sum_{j=1}^{p} \mathbf{x}_j \mu_{blj}\alpha_{lj}$

**for** $l = 1, \cdots, L$ **do**

$\tilde{\boldsymbol{r}}_l \leftarrow \tilde{\boldsymbol{r}} + \mathbf{X}^T \sum_{j=1}^{p} \mathbf{x}_j \mu_{blj} \alpha_{lj}$

$\boldsymbol{\Sigma}_{wl}^{-1} \leftarrow \mathbf{I}_M / \sigma_{wl}^2 + 2 \sum_{j=1}^{p} \lambda(\xi_{lj}) \boldsymbol{A}_j \boldsymbol{A}_j^T$

$\boldsymbol{\mu}_{wl} \leftarrow \boldsymbol{\Sigma}_{wl} \sum_{j=1}^{p} (\alpha_{lj} - 1/2 + 2\rho_l \lambda(\xi_{lj})) \boldsymbol{A}_j$

$s_{blj}^2 \leftarrow 1/(\boldsymbol{x}_j^T \boldsymbol{x}_j / \sigma^2 + 1/\sigma_{bl}^2), j = 1, \cdots, p$

$\mu_{blj} \leftarrow s_{blj}^2 \cdot \tilde{\boldsymbol{r}}_{lj} / \sigma^2, j = 1, \cdots, p$, where $\tilde{\boldsymbol{r}}_{lj}$ is the $j$-th coordinate element of $\tilde{\boldsymbol{r}}_l$

$\xi_{lj}^2 \leftarrow \boldsymbol{A}_j^T (\boldsymbol{\Sigma}_{wl} + \boldsymbol{\mu}_{wl} \boldsymbol{\mu}_{wl}^T) \boldsymbol{A}_j + \rho_l^2 - 2\rho_l \boldsymbol{A}_j^T \boldsymbol{\mu}_l, j = 1, \cdots, p$

$\rho_l \leftarrow [(p/2 - 1)/2 + \sum_{j=1}^{p} \lambda(\xi_{lj}) \boldsymbol{A}_j^T \boldsymbol{\mu}_{wl}] / \sum_{j=1}^{p} \lambda(\xi_{lj})$

$\sigma_{wl}^2 \leftarrow \mathrm{Tr}(\boldsymbol{\Sigma}_{wl} + \boldsymbol{\mu}_{wl} \boldsymbol{\mu}_{wl}^T)/m$

$\sigma_{bl}^2 \leftarrow \sum_{j=1}^{p} \alpha_{lj} (\mu_{blj}^2 + s_{blj}^2)$

$\tilde{\boldsymbol{r}} \leftarrow \tilde{\boldsymbol{r}}_l - \mathbf{X}^T \sum_{j=1}^{p} \mathbf{x}_j \mu_{blj} \alpha_{lj}$

**end for**

$\sigma^2 \leftarrow \mathrm{ERSS}/n$, where ERSS is given by (35)

**until** convergence criterion satisfied

**Output:** $\sigma^2$, $\{\sigma_{bl}^2\}$, $\{\sigma_{wl}^2\}$ $\{\mu_{blj}\}$, $\{s_{blj}^2\}$, $\{\boldsymbol{\mu}_{wl}\}$, $\{\boldsymbol{\Sigma}_{wl}\}$, $\{\xi_{lj}\}$, $\{\rho_l\}$

#### 2.3 Stage 3: Funmap

**Algorithm 3** IBSS algorithm for Funmap

**Input:** Data $\mathbf{X}^T\mathbf{X}$, $\mathbf{X}^T\mathbf{y}$, $\mathbf{y}^T\mathbf{y}$ (which can be calculated from $\mathbf{z}$, $\mathbf{R}$ and $n$) and $\boldsymbol{A}$

**Input:** Initial value $\{\alpha_{lj}\}$ from the Output of Stage 1

**Input:** Initial value $\sigma^2$, $\{\sigma_{bl}^2\}$, $\{\sigma_{wl}^2\}$ $\{\mu_{blj}\}$, $\{s_{blj}^2\}$, $\{\boldsymbol{\mu}_{wl}\}$, $\{\boldsymbol{\Sigma}_{wl}\}$, $\{\xi_{lj}\}$, $\{\rho_l\}$ from Stage 2

Initialize: $\sigma^2$, $\{\sigma_{bl}^2\}$, $\{\sigma_{wl}^2\}$ $\{\mu_{blj}\}$, $\{s_{blj}^2\}$, $\{\boldsymbol{\mu}_{wl}\}$, $\{\boldsymbol{\Sigma}_{wl}\}$, $\{\xi_{lj}\}$, $\{\rho_l\}$ and $\{\alpha_{lj}\}$

**repeat**

$\tilde{\boldsymbol{r}} \leftarrow \mathbf{X}^T \boldsymbol{y} - \sum_{l=1}^{L} \mathbf{X}^T \sum_{j=1}^{p} \mathbf{x}_j \mu_{blj} \alpha_{lj}$

**for** $l = 1, \cdots, L$ **do**

$\tilde{\boldsymbol{r}}_l \leftarrow \tilde{\boldsymbol{r}} + \mathbf{X}^T \sum_{j=1}^{p} \mathbf{x}_j \mu_{blj} \alpha_{lj}$

$\boldsymbol{\Sigma}_{wl}^{-1} \leftarrow \mathbf{I}_M / \sigma_{wl}^2 + 2 \sum_{j=1}^{p} \lambda(\xi_{lj}) \boldsymbol{A}_j \boldsymbol{A}_j^T$

$\boldsymbol{\mu}_{wl} \leftarrow \boldsymbol{\Sigma}_{wl} \sum_{j=1}^{p} (\alpha_{lj} - 1/2 + 2\rho_l \lambda(\xi_{lj})) \boldsymbol{A}_j$

$s_{blj}^2 \leftarrow 1/(\boldsymbol{x}_j^T \boldsymbol{x}_j / \sigma^2 + 1/\sigma_{bl}^2), j = 1, \cdots, p$

$\mu_{blj} \leftarrow s_{blj}^2 \cdot \tilde{\boldsymbol{r}}_{lj} / \sigma^2, j = 1, \cdots, p$, where $\tilde{\boldsymbol{r}}_{lj}$ is the $j$-th coordinate element of $\tilde{\boldsymbol{r}}_l$

$\xi_{lj}^2 \leftarrow \boldsymbol{A}_j^T (\boldsymbol{\Sigma}_{wl} + \boldsymbol{\mu}_{wl} \boldsymbol{\mu}_{wl}^T) \boldsymbol{A}_j + \rho_l^2 - 2\rho_l \boldsymbol{A}_j^T \boldsymbol{\mu}_l, j = 1, \cdots, p$

$\rho_l \leftarrow [(p/2 - 1)/2 + \sum_{j=1}^{p} \lambda(\xi_{lj}) \boldsymbol{A}_j^T \boldsymbol{\mu}_{wl}] / \sum_{j=1}^{p} \lambda(\xi_{lj})$

$\alpha_{lj} \leftarrow \exp(u_j) / \sum_{j'=1}^{p} \exp(u_{j'}), \text{where } u_j = \boldsymbol{A}_j^T \boldsymbol{\mu}_{wl} + \dfrac{\mu_{blj}^2}{2 s_{blj}^2} + \dfrac{1}{2} \log s_{blj}^2, j = 1, \cdots, p$

$\sigma_{wl}^2 \leftarrow \mathrm{Tr}(\boldsymbol{\Sigma}_{wl} + \boldsymbol{\mu}_{wl} \boldsymbol{\mu}_{wl}^T)/m$

$\sigma_{bl}^2 \leftarrow \sum_{j=1}^{p} \alpha_{lj} (\mu_{blj}^2 + s_{blj}^2)$

$\tilde{\boldsymbol{r}} \leftarrow \tilde{\boldsymbol{r}}_l - \mathbf{X}^T \sum_{j=1}^{p} \mathbf{x}_j \mu_{blj} \alpha_{lj}$

**end for**

$\sigma^2 \leftarrow \mathrm{ERSS}/n$, where ERSS is given by (35)

**until** convergence criterion satisfied

**Output:** $\sigma^2$, $\{\sigma_{bl}^2\}$, $\{\sigma_{wl}^2\}$ $\{\mu_{blj}\}$, $\{s_{blj}^2\}$, $\{\boldsymbol{\mu}_{wl}\}$, $\{\boldsymbol{\Sigma}_{wl}\}$, $\{\xi_{lj}\}$, $\{\rho_l\}$

### 3 More simulation results

#### 3.1 Comparison of results with different sample sizes

We set the number of casual SNPs $L_0 = 2, 3$ and generated simulation data with different sample size $n = 20000, 50000$. The results are shown in Figure S1-S12. All the results are summarized from 500 replications across 10 gene regions.

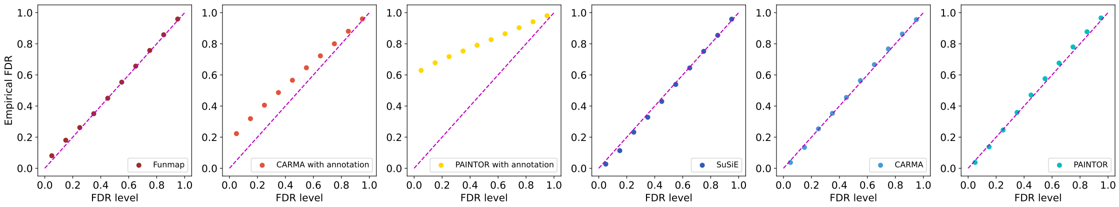

Figure S1: Comparison of empirical FDR with $n = 20000, m = 100, L_0 = 2, \phi = 0.0075$.

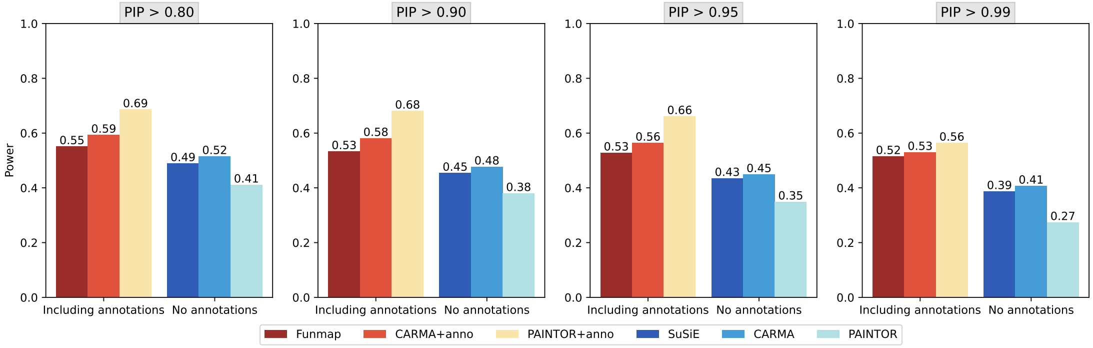

Figure S2: Comparison of statistical power with $n = 20000, m = 100, L_0 = 2, \phi = 0.0075$.

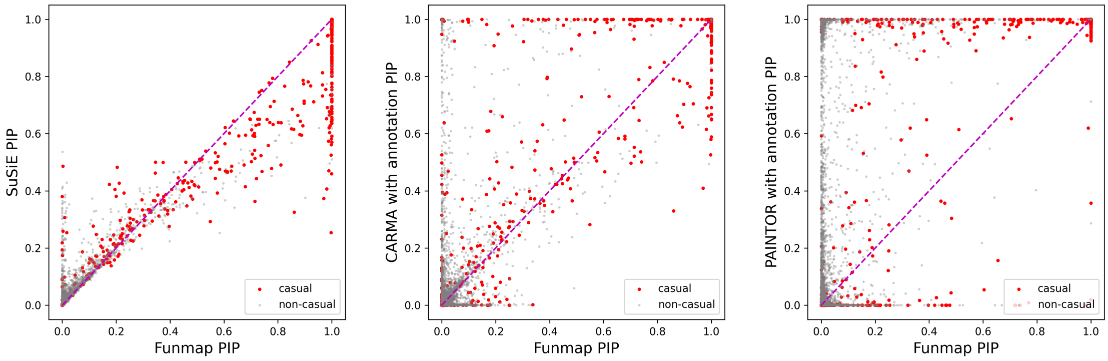

Figure S3: Comparison of PIP scatter with $n = 20000, m = 100, L_0 = 2, \phi = 0.0075$.

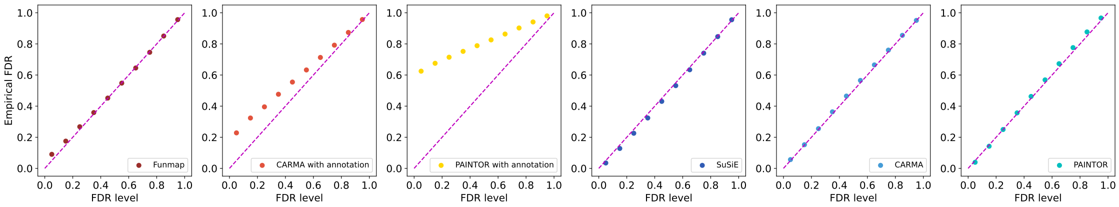

Figure S4: Comparison of empirical FDR with $n = 20000, m = 100, L_0 = 3, \phi = 0.0075$.

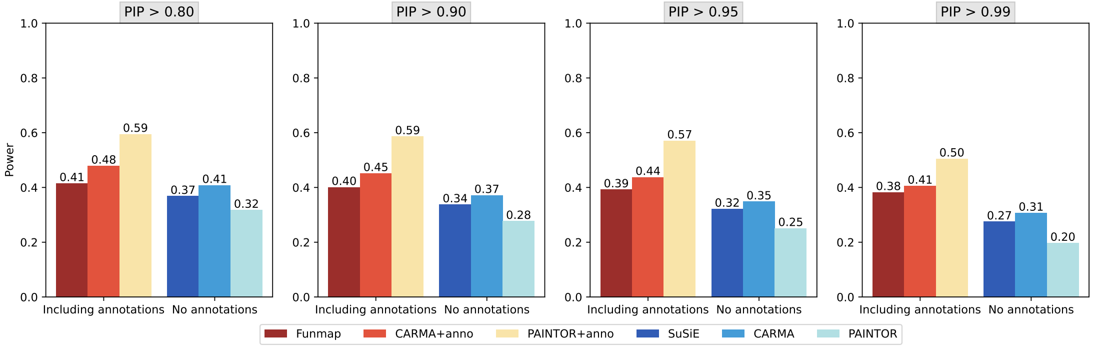

Figure S5: Comparison of statistical power with $n = 20000, m = 100, L_0 = 3, \phi = 0.0075$.

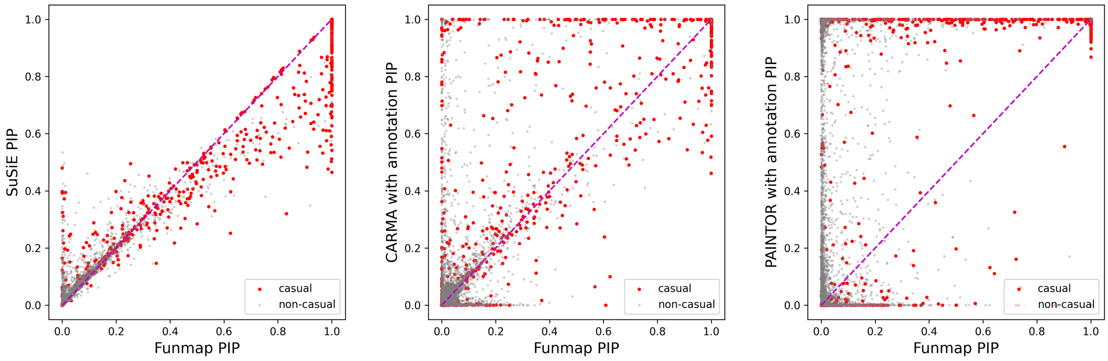

Figure S6: Comparison of PIP scatter with $n = 20000, m = 100, L_0 = 3, \phi = 0.0075$.

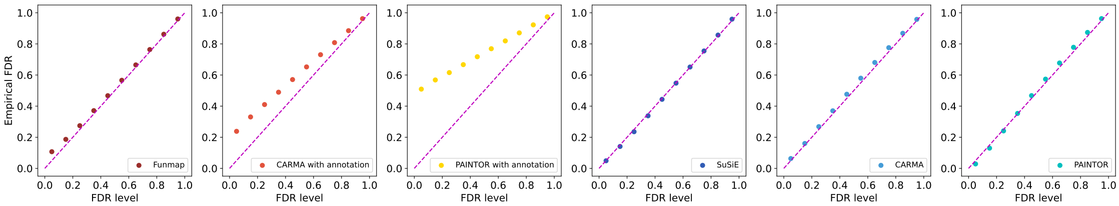

Figure S7: Comparison of empirical FDR with $n = 50000, m = 100, L_0 = 2, \phi = 0.0075$.

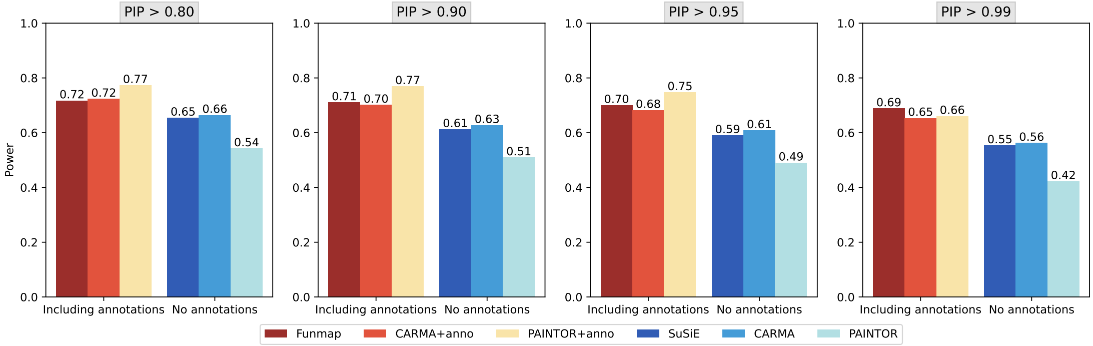

Figure S8: Comparison of statistical power with $n = 50000, m = 100, L_0 = 2, \phi = 0.0075$.

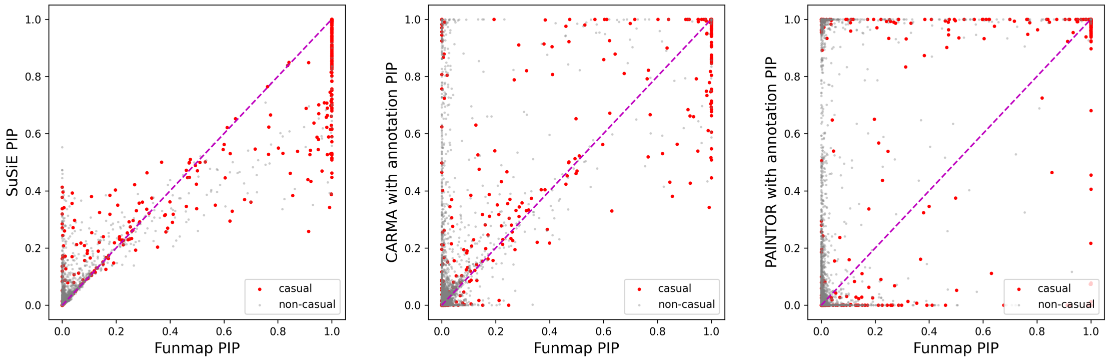

Figure S9: Comparison of PIP scatter with $n = 50000, m = 100, L_0 = 2, \phi = 0.0075$.

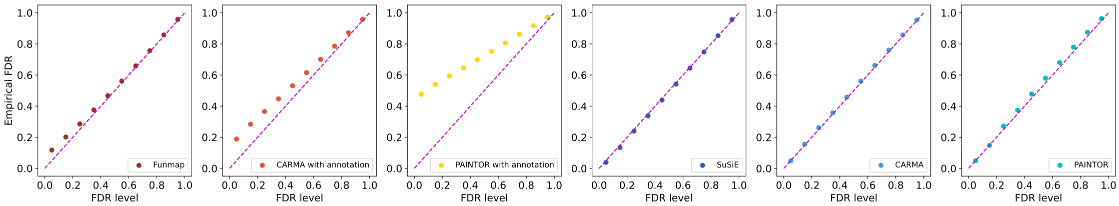

Figure S10: Comparison of empirical FDR with $n = 50000, m = 100, L_0 = 3, \phi = 0.0075$.

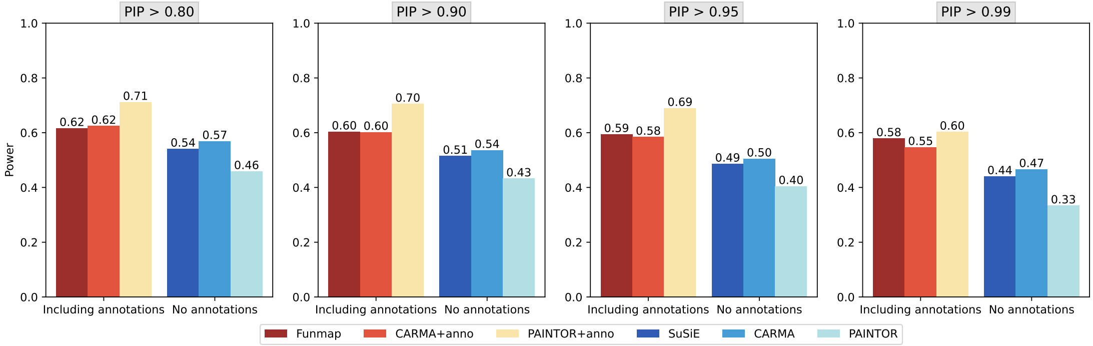

Figure S11: Comparison of statistical power with $n = 50000, m = 100, L_0 = 3, \phi = 0.0075$.

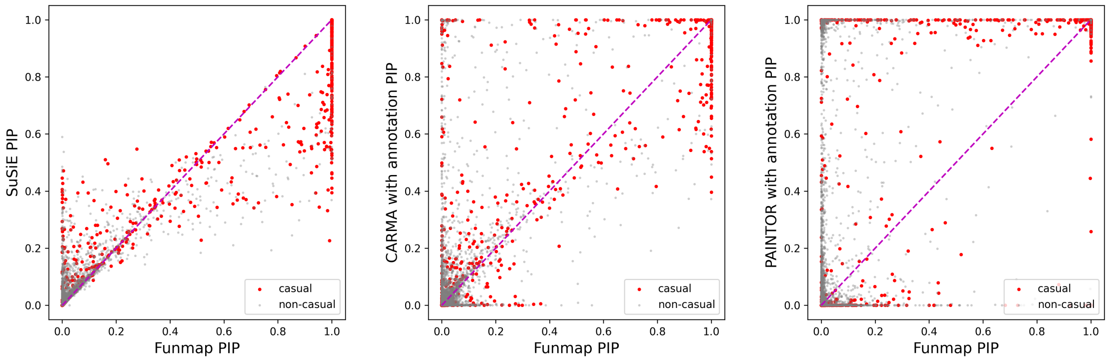

Figure S12: Comparison of PIP scatter with $n = 50000, m = 100, L_0 = 3, \phi = 0.0075$.

#### 3.2 Comparison of results with different number of annotations

We set the number of casual SNPs $L_0 = 2, 3$ and generated simulation data with different annotation numbers $m = 100, 50, 20$. The results are shown in Figure S13-S30. All the results are summarized from the summation of 500 replications from 10 gene regions.

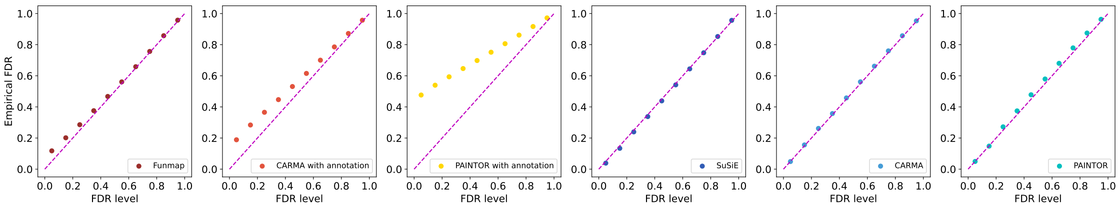

Figure S13: Comparison of empirical FDR with $n = 50000, m = 100, L_0 = 3, \phi = 0.0075$.

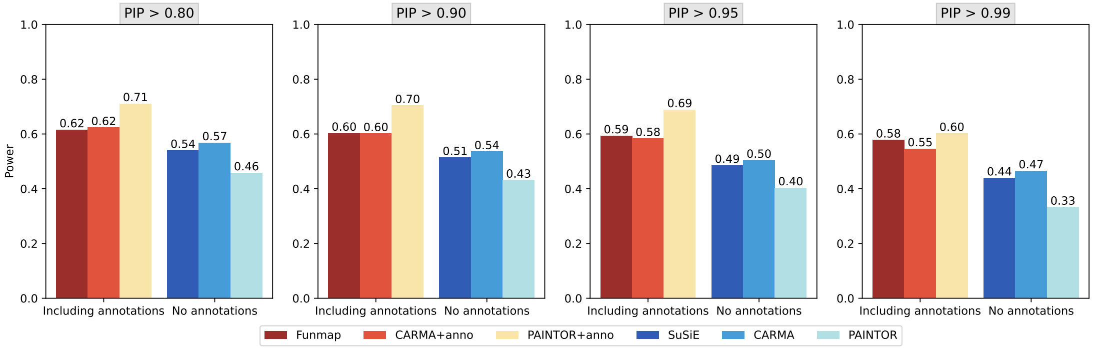

Figure S14: Comparison of statistical power with $n = 50000, m = 100, L_0 = 3, \phi = 0.0075$.

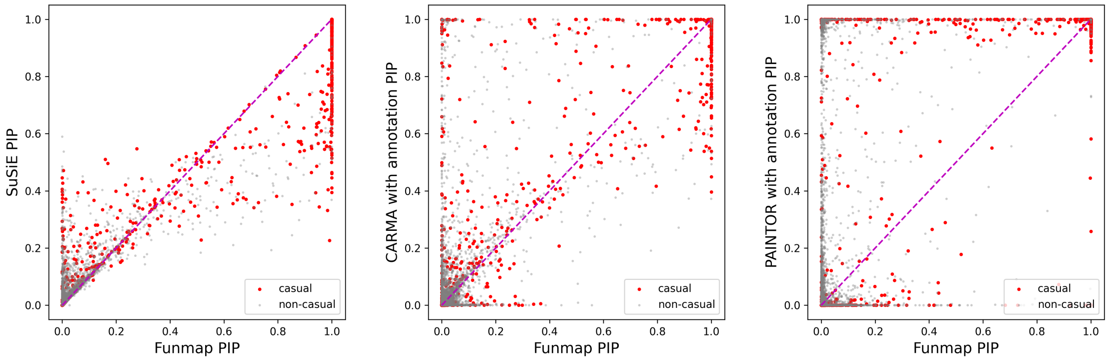

Figure S15: Comparison of PIP scatter with $n = 50000, m = 100, L_0 = 3, \phi = 0.0075$.

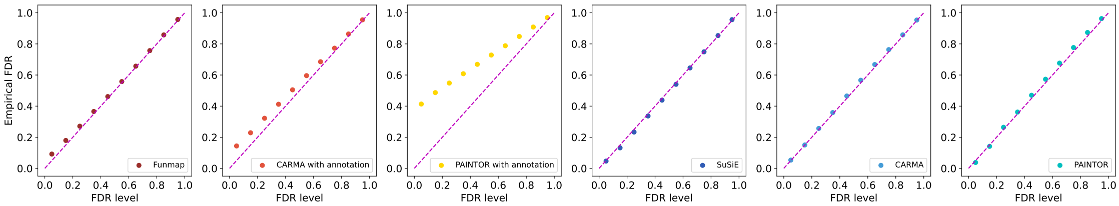

Figure S16: Comparison of empirical FDR with $n = 50000, m = 50, L_0 = 3, \phi = 0.0075$.

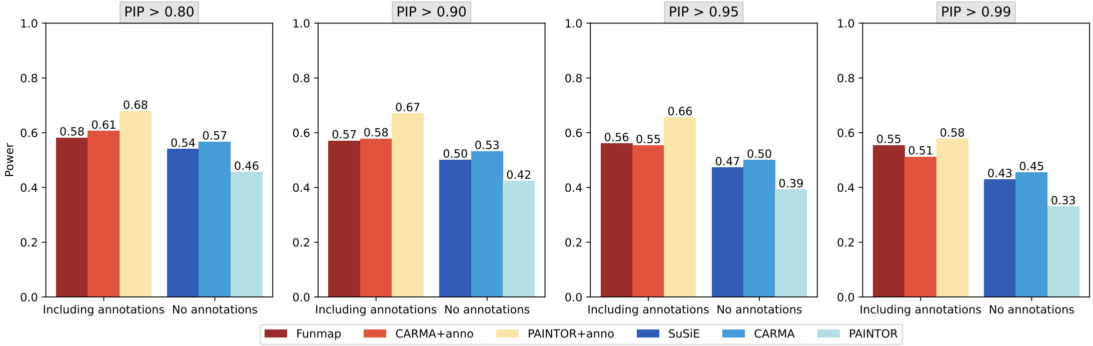

Figure S17: Comparison of statistical power with $n = 50000, m = 50, L_0 = 3, \phi = 0.0075$.

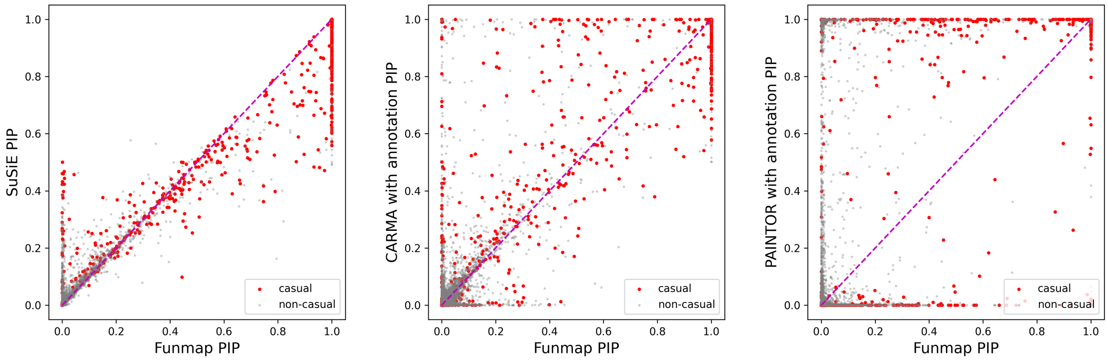

Figure S18: Comparison of PIP scatter with $n = 50000, m = 50, L_0 = 3, \phi = 0.0075$.

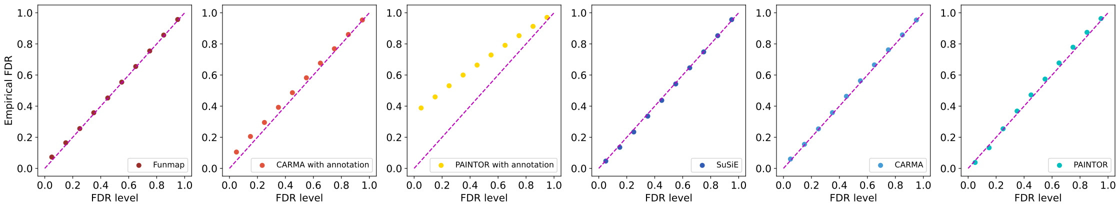

Figure S19: Comparison of empirical FDR with $n = 50000, m = 20, L_0 = 3, \phi = 0.0075$.

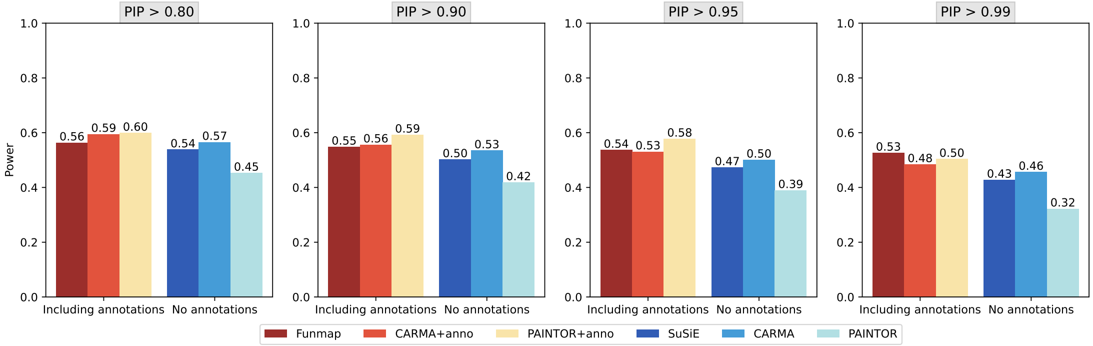

Figure S20: Comparison of statistical power with $n = 50000, m = 20, L_0 = 3, \phi = 0.0075$.

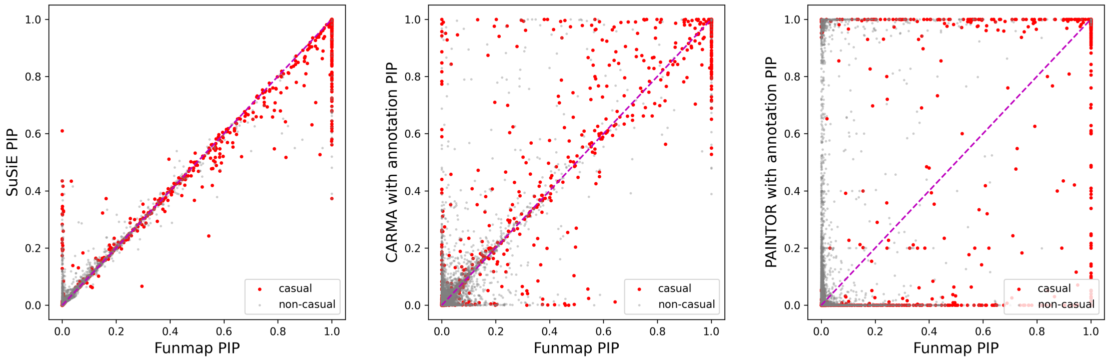

Figure S21: Comparison of PIP scatter with $n = 50000, m = 20, L_0 = 3, \phi = 0.0075$.

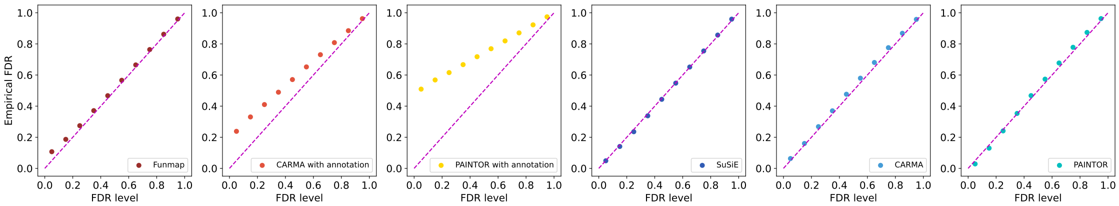

Figure S22: Comparison of empirical FDR with $n = 50000, m = 100, L_0 = 2, \phi = 0.0075$.

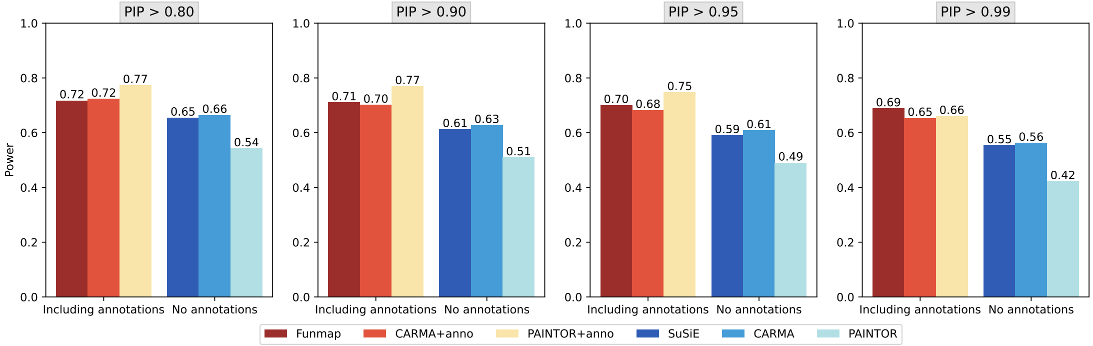

Figure S23: Comparison of statistical power with $n = 50000, m = 100, L_0 = 2, \phi = 0.0075$.

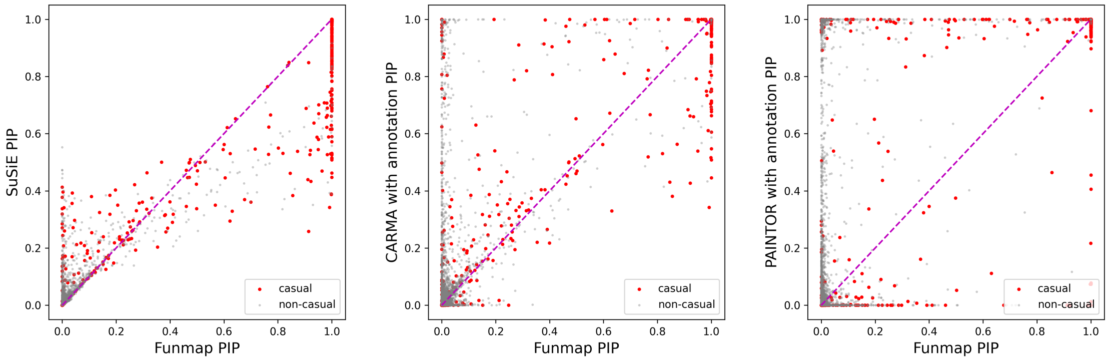

Figure S24: Comparison of PIP scatter with $n = 50000, m = 100, L_0 = 2, \phi = 0.0075$.

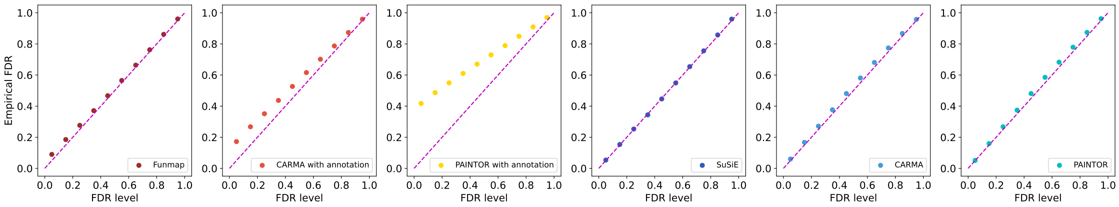

Figure S25: Comparison of empirical FDR with $n = 50000, m = 50, L_0 = 2, \phi = 0.0075$.

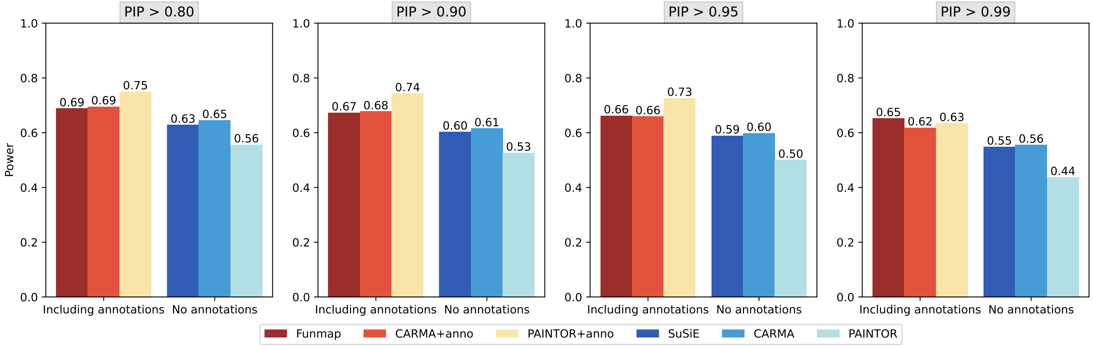

Figure S26: Comparison of statistical power with $n = 50000, m = 50, L_0 = 2, \phi = 0.0075$.

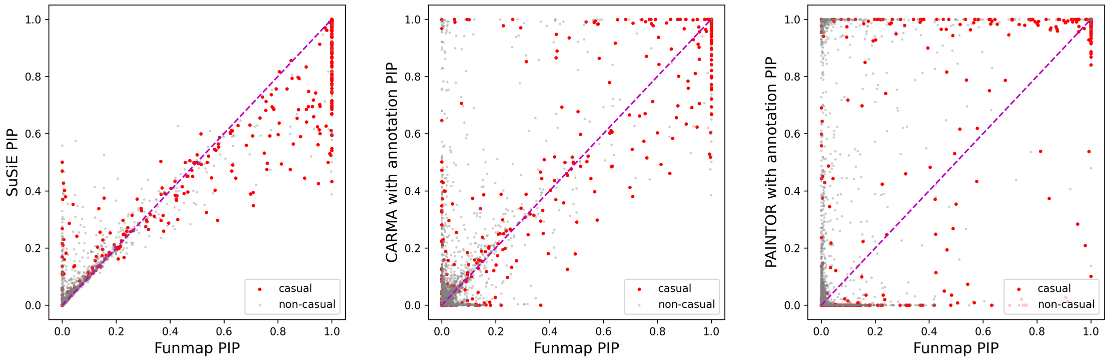

Figure S27: Comparison of PIP scatter with $n = 50000, m = 50, L_0 = 2, \phi = 0.0075$.

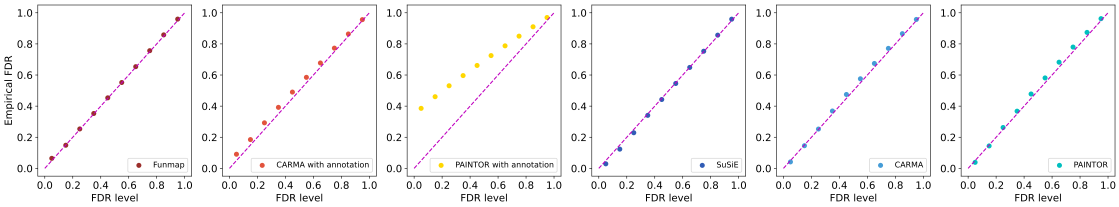

Figure S28: Comparison of empirical FDR with $n = 50000, m = 20, L_0 = 2, \phi = 0.0075$.

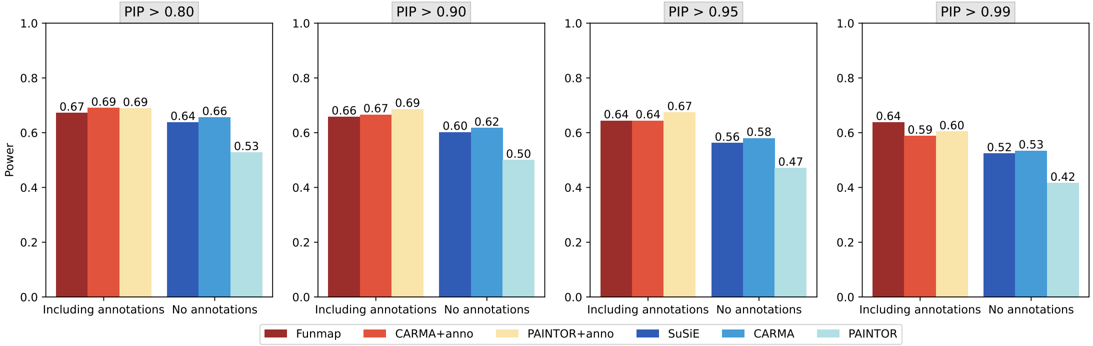

Figure S29: Comparison of statistical power with $n = 50000, m = 20, L_0 = 2, \phi = 0.0075$.

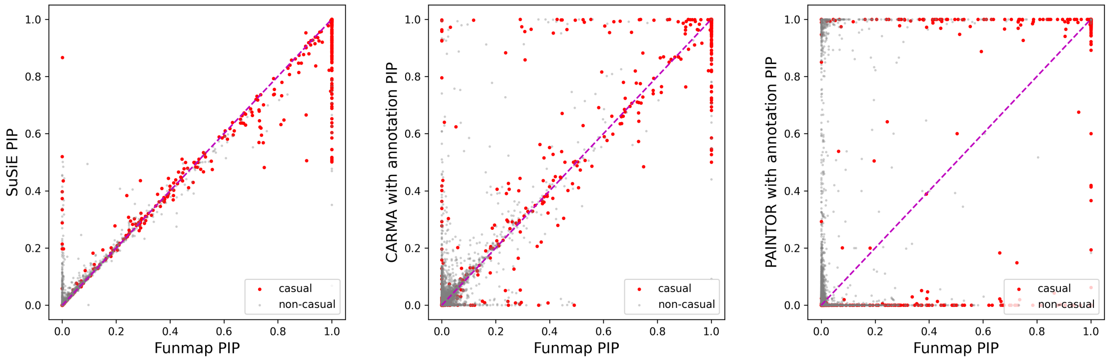

Figure S30: Comparison of PIP scatter with $n = 50000, m = 20, L_0 = 2, \phi = 0.0075$.

#### 3.3 Comparison of results with sparse annotations

We set the number of casual SNPs $L_0 = 2, 3$ and generated simulation data with different annotation numbers $m = 100, 50, 20$, but only half of the annotations are truly informative. The results are shown in Figure S31-S48. All the results are summarized from the summation of 500 replications from 10 gene regions.

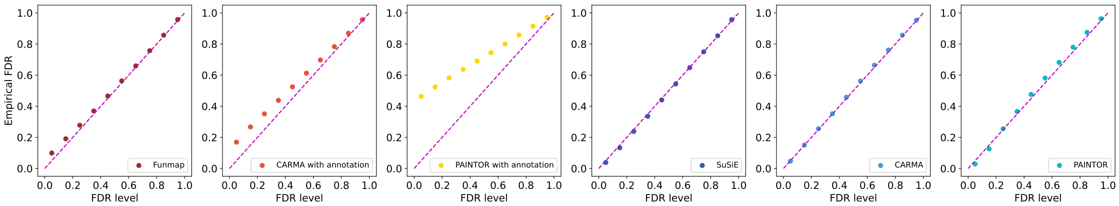

Figure S31: Comparison of empirical FDR with $n = 50000, m = 100, m_{anno} = 50, L_0 = 3$.

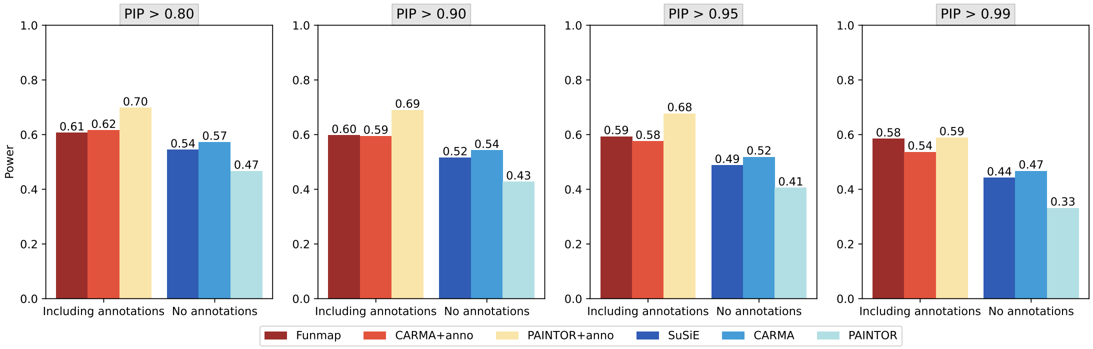

Figure S32: Comparison of statistical power with $n = 50000, m = 100, m_{anno} = 50, L_0 = 3$.

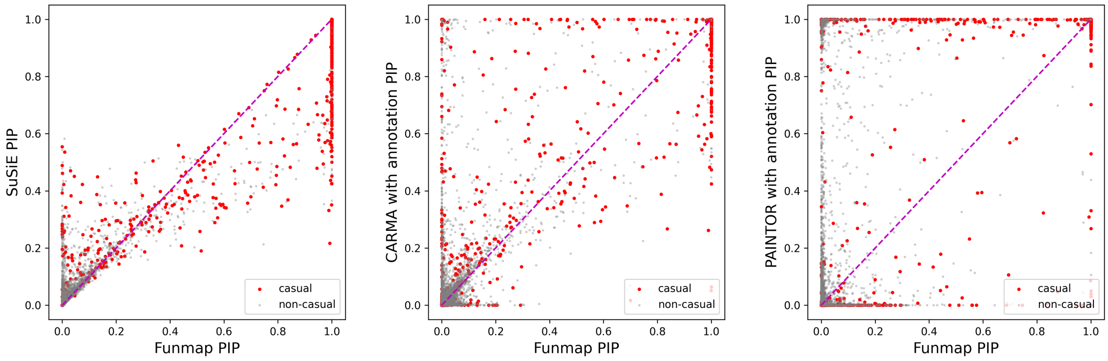

Figure S33: Comparison of PIP scatter with $n = 50000, m = 100, m_{anno} = 50, L_0 = 3$.

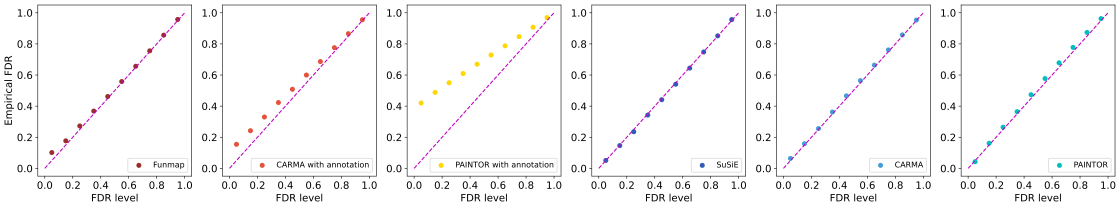

Figure S34: Comparison of empirical FDR with $n = 50000, m = 50, m_{anno} = 25, L_0 = 3$.

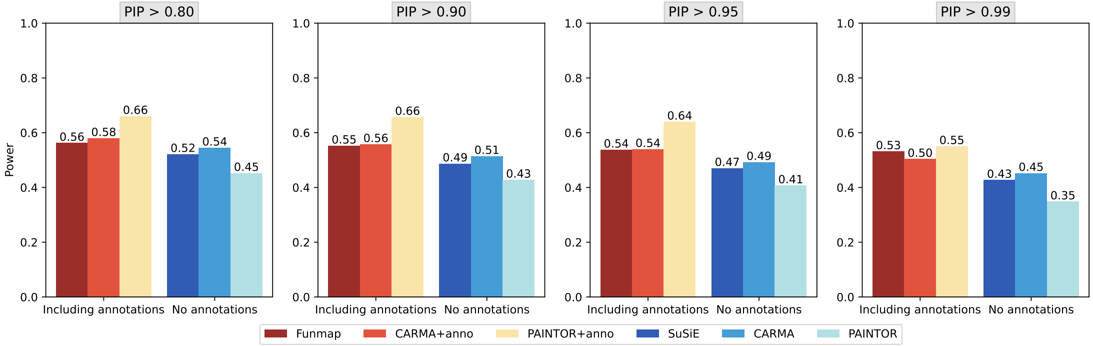

Figure S35: Comparison of statistical power with $n = 50000, m = 50, m_{anno} = 25, L_0 = 3$.

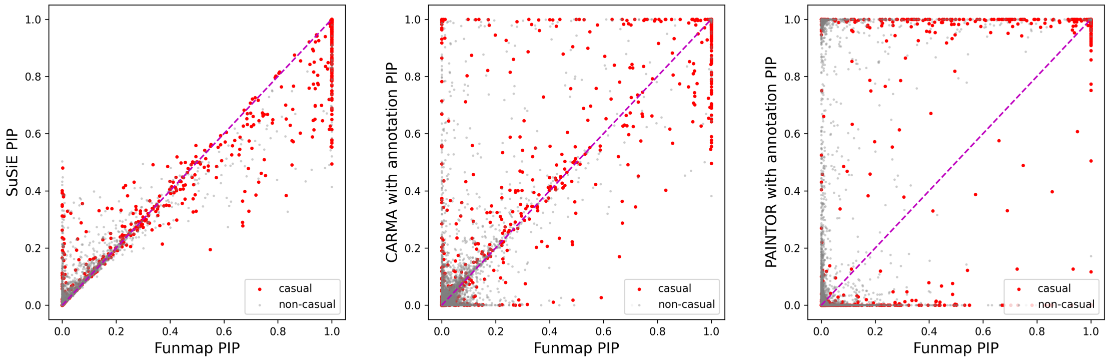

Figure S36: Comparison of PIP scatter with $n = 50000, m = 50, m_{anno} = 25, L_0 = 3$.

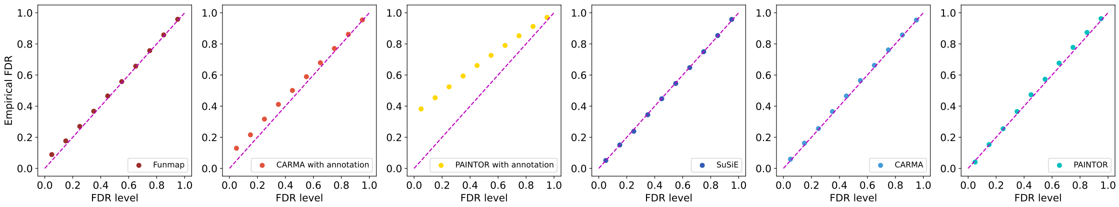

Figure S37: Comparison of empirical FDR with $n = 50000, m = 20, m_{anno} = 10, L_0 = 3$.

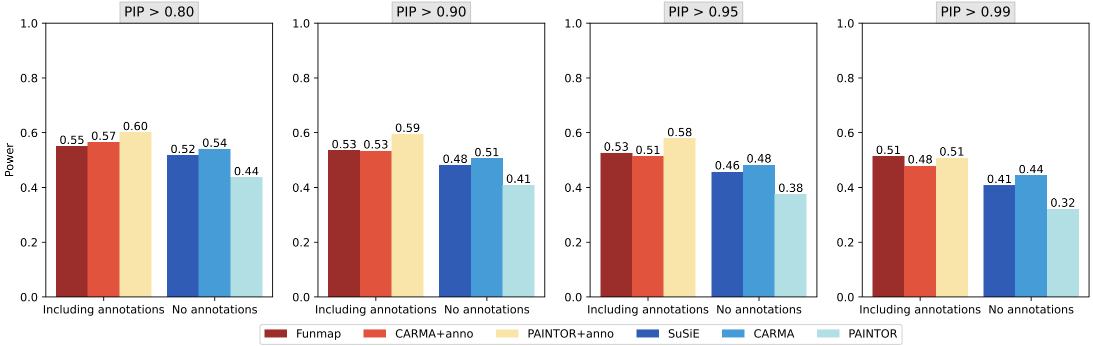

Figure S38: Comparison of statistical power with $n = 50000, m = 20, m_{anno} = 10, L_0 = 3$.

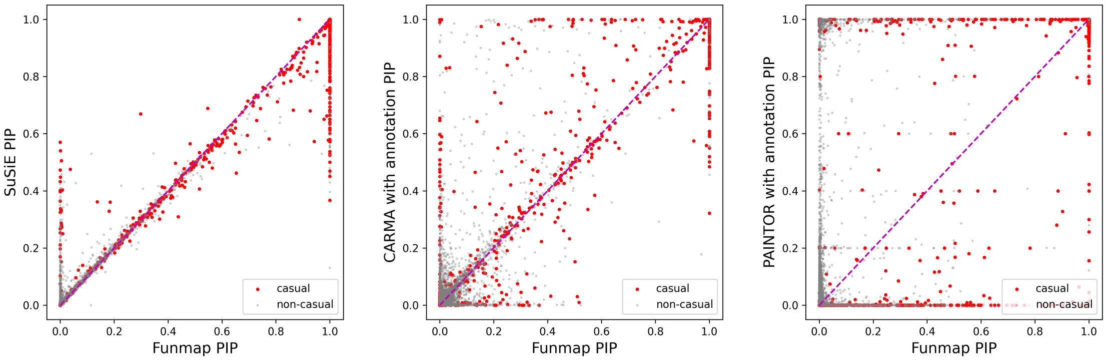

Figure S39: Comparison of PIP scatter with $n = 50000, m = 20, m_{anno} = 10, L_0 = 3$.

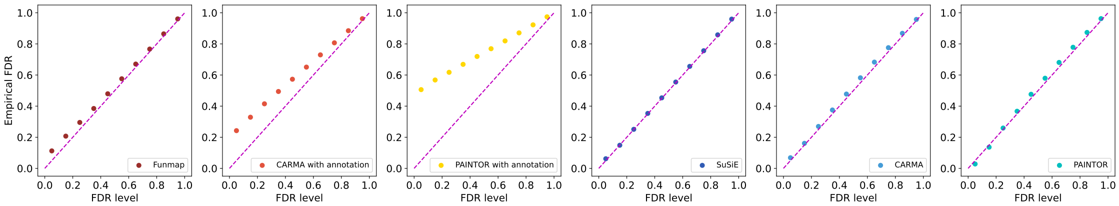

Figure S40: Comparison of empirical FDR with $n = 50000, m = 100, m_{anno} = 50, L_0 = 2$.

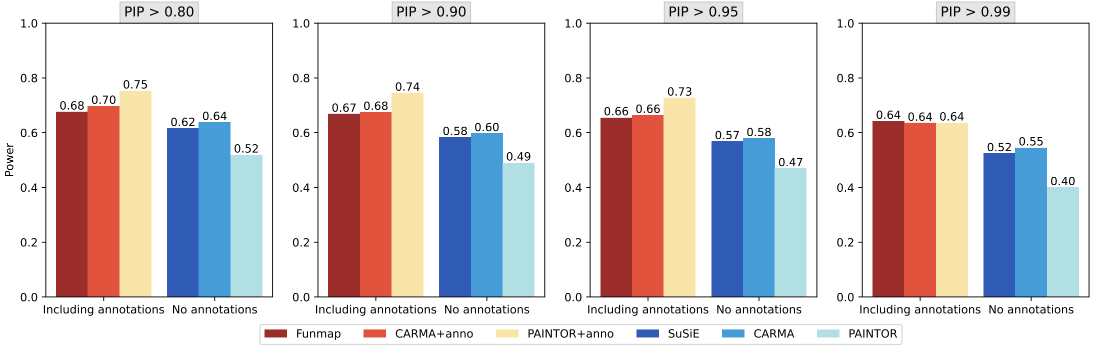

Figure S41: Comparison of statistical power with $n = 50000, m = 100, m_{anno} = 50, L_0 = 2$.

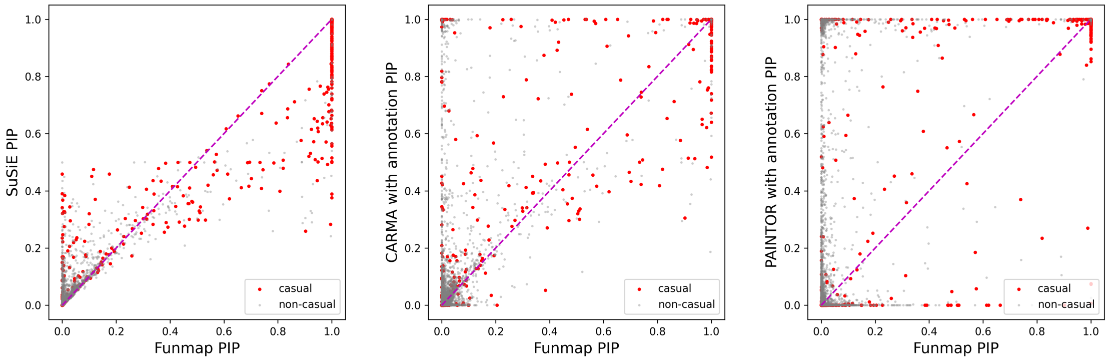

Figure S42: Comparison of PIP scatter with $n = 50000, m = 100, m_{anno} = 50, L_0 = 2$.

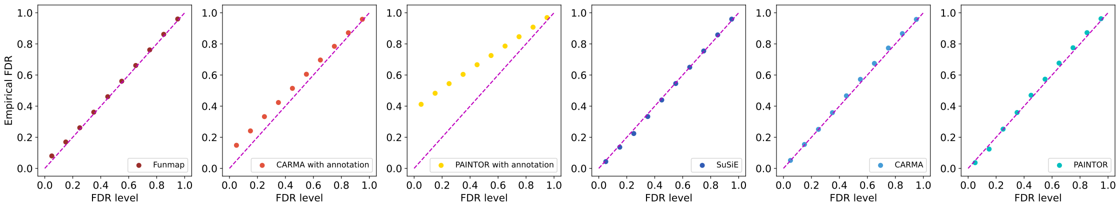

Figure S43: Comparison of empirical FDR with $n = 50000, m = 50, m_{anno} = 25, L_0 = 2$.

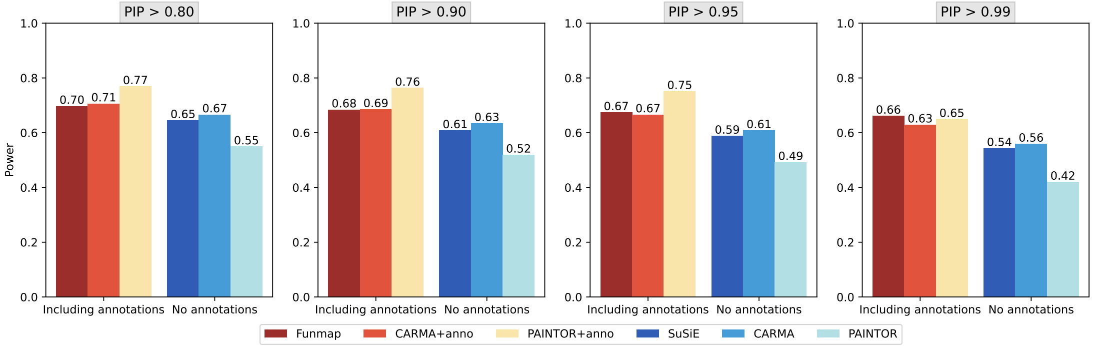

Figure S44: Comparison of statistical power with $n = 50000, m = 50, m_{anno} = 25, L_0 = 2$.

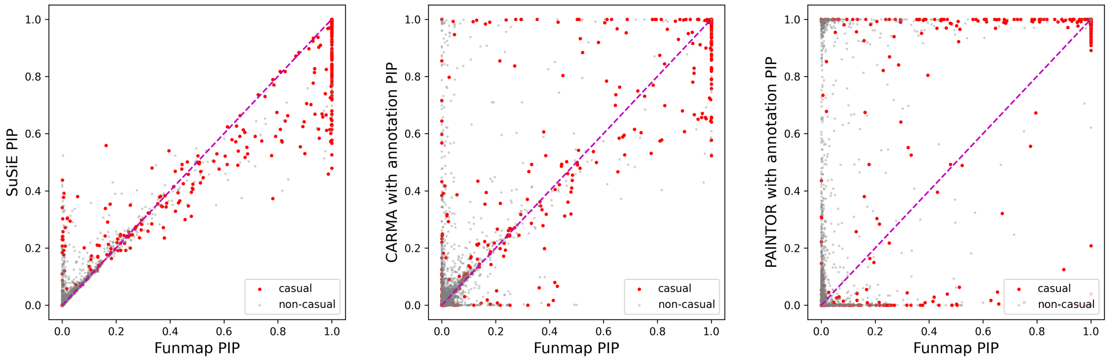

Figure S45: Comparison of PIP scatter with $n = 50000, m = 50, m_{anno} = 25, L_0 = 2$.

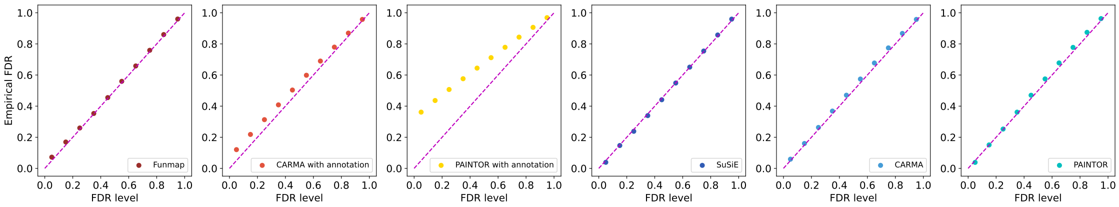

Figure S46: Comparison of empirical FDR with $n = 50000, m = 20, m_{anno} = 10, L_0 = 2$.

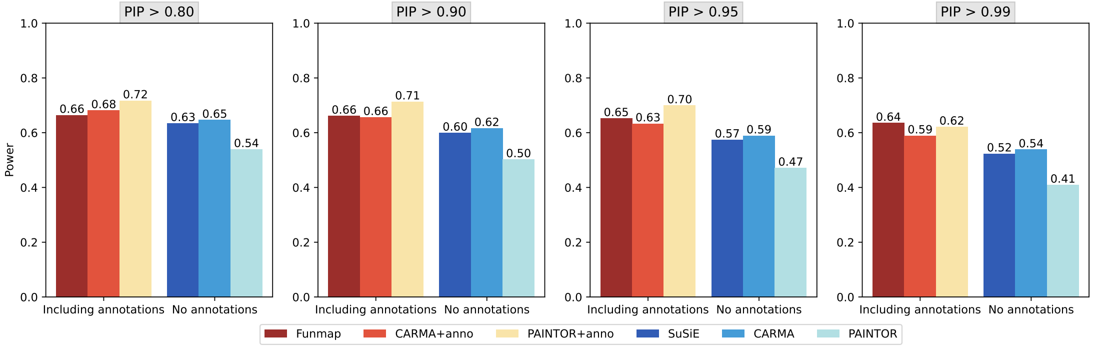

Figure S47: Comparison of statistical power with $n = 50000, m = 20, m_{anno} = 10, L_0 = 2$.

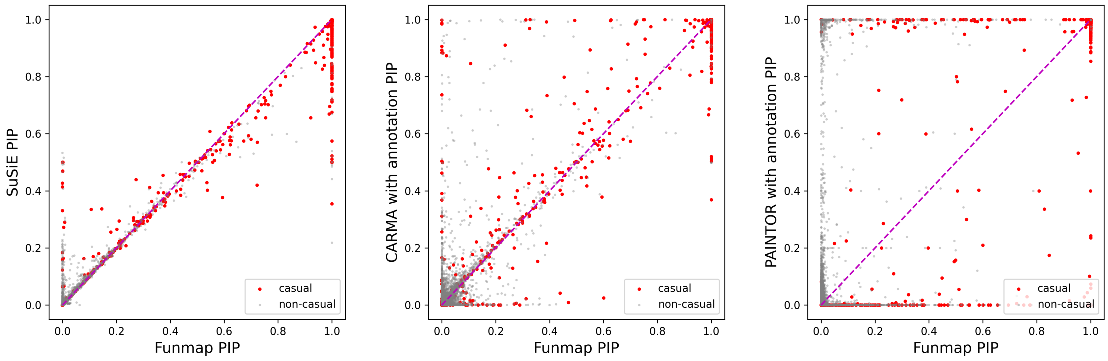

Figure S48: Comparison of PIP scatter with $n = 50000, m = 20, m_{anno} = 10, L_0 = 2$.

#### 4 Supplementry table

Table S1: Simulation loci

| gene name | Chromosome | region start | region end | SNP numbers |
|---|---|---|---|---|
| TNNI1 | chr1 | 200937832 | 201937832 | 1272 |
| ALS2CR12 | chr2 | 201681247 | 202681247 | 712 |
| IGFBP5 | chr2 | 217405832 | 218796508 | 1578 |
| | chr5 | 44013304 | 45206498 | 1313 |
| MAP3K1 | chr5 | 55531884 | 56587883 | 854 |
| C6orf211 | chr6 | 151418856 | 152937016 | 2561 |
| | chr7 | 130167121 | 131167121 | 854 |
| MYC | chr8 | 127424659 | 130041931 | 4107 |
| FGFR2 | chr10 | 122593901 | 123849324 | 1897 |
| TBX3 | chr12 | 115336522 | 116336522 | 1833 |

Table S2: The number of putative causal SNPs under different PIP thresholds.

| | HDL | | | LDL | | | TG | | | TC | | |
|---|---|---|---|---|---|---|---|---|---|---|---|---|
| PIP Threshold | 0.90 | 0.95 | 0.99 | 0.90 | 0.95 | 0.99 | 0.90 | 0.95 | 0.99 | 0.90 | 0.95 | 0.99 |
| Funmap | 583 | 574 | 566 | 308 | 304 | 298 | 493 | 486 | 477 | 357 | 347 | 342 |
| CARMA+anno | 1056 | 962 | 762 | 538 | 493 | 411 | 959 | 862 | 679 | 660 | 614 | 486 |
| PAINTOR+anno | 1715 | 1255 | 527 | 859 | 601 | 229 | 1480 | 1063 | 413 | 951 | 683 | 239 |
| SuSiE | 167 | 148 | 115 | 90 | 81 | 62 | 116 | 100 | 81 | 110 | 96 | 74 |
| CARMA | 326 | 281 | 203 | 184 | 151 | 117 | 263 | 222 | 160 | 209 | 188 | 147 |
| PAINTOR | 142 | 100 | 36 | 88 | 61 | 16 | 94 | 62 | 25 | 71 | 51 | 24 |

#### 5 Code and data availability

Fummap is available at https://github.com/LeeHITsz/Funmap

SuSiE is available at https://github.com/stephenslab/susieR

CARMA is available at https://github.com/ZikunY/CARMA

PAINTOR (3.0) is available at https://github.com/gkichaev/PAINTOR_V3.0

Plink is available at https://www.cog-genomics.org/plink

Selected gene regions in simulation are available at https://github.com/ZikunY/CARMA/tree/master/Simulation%20Study

Summary data of Lipid-related traits are available at https://nealelab.github.io/UKBB_ldsc/index.html

LD files for UKBB British-ancestry are available at https://alkesgroup.broadinstitute.org/UKBB_LD.ThescATAC-seqdata

Functional annotations for real data are available at https://alkesgroup.broadinstitute.org/LDSCORE/baselineLD_v2.1_annots

Summary statistics from GLGC can be downloaded at https://csg.sph.umich.edu/willer/public/glgc-lipids2021

#### References

[1] Gao Wang, Abhishek Sarkar, Peter Carbonetto, and Matthew Stephens. A simple new approach to variable selection in regression, with application to genetic fine mapping. *Journal of the Royal Statistical Society Series B: Statistical Methodology*, 82(5):1273–1300, 2020.

[2] Guillaume Bouchard. Efficient bounds for the softmax function and applications to approximate inference in hybrid models. In *NIPS 2007 workshop for approximate Bayesian inference in continuous/hybrid systems*, volume 6, 2007.

[3] Yongtao Guan and Matthew Stephens. Bayesian variable selection regression for genome-wide association studies and other large-scale problems. *The Annals of Applied Statistics*, 5(3):1780–1815, 2011.

[4] Peter Carbonetto and Matthew Stephens. Scalable variational inference for bayesian variable selection in regression, and its accuracy in genetic association studies. *Bayesian analysis*, 7(1):73–108, 2012.

[5] Yuxin Zou, Peter Carbonetto, Gao Wang, and Matthew Stephens. Fine-mapping from summary data with the "sum of single effects" model. *PLoS Genetics*, 18(7):e1010299, 2022.
